# Recreational Use of Pregabalin in Ukraine. Availability, Patterns of Use and Chemsex

**DOI:** 10.1101/2025.10.06.25337361

**Authors:** Mykyta Lapin, Maryna Shevchenko

## Abstract

**Introduction:** Substance use disorders pose significant public health challenges and are associated with a wide range of social and medical consequences. Pregabalin, a gabapentinoid, was approved by the U.S. Food and Drug Administration in 2004 for the treatment of specific medical conditions. Despite its initially limited indications, the drug achieved substantial commercial success, ranking among the top ten global pharmaceutical sales by 2017. Reports of addictive potential of pregabalin began to surface shortly after its market release in 2005. The first documented cases of misuse appeared in Europe in 2008, and by 2010, the clinical community had begun to recognize pregabalin use disorder. Since then, the prevalence of pregabalin misuse has increased across all age groups globally. Its recreational use contributes to the overall burden of both communicable and non-communicable diseases, intensifying existing public health threats. This study examines the phenomenon of recreational pregabalin use in Ukraine. The research focuses on its impact on the physical and mental health of users, as well as on their socio-economic well-being. The primary aim is to explore existing patterns of recreational use to inform harm reduction strategies and public health interventions.

**Methods:** The inclusion criterion for the study was recreational use of pregabalin at the time of the study or a history of such use. The sample was formed by the snowball sampling method, data collection was conducted through semi-structured interviews, and analysis was performed using the phenomenological method. The Drugs Wheel model was used to classify psychoactive substances.

**Results:** Substance users are not aware of the risks of recreational use of pregabalin. Pregabalin is available for recreational use in Ukraine through over-the-counter pharmacies. Patterns of recreational use of pregabalin in Ukraine are dangerous in terms of mental and physical health disorders and negative socioeconomic consequences. Pregabalin is used as a tool for chemsex, and users neglect harm reduction measures for risky sexual behavior during such contacts. The difficulty of quitting recreational pregabalin use has been documented, and adherence to the existing mental health care system is low.

**Discussion:** Low awareness of the risks of recreational use of pregabalin among substance users creates a need for educational activities among target groups. Given the commitment of doctors to prescribing pregabalin and the lack of warnings about the negative consequences, it is imperative to educate healthcare professionals about the risks of such prescriptions. Since pregabalin is used as a tool for chemsex, and users neglect harm reduction measures for such contacts, it is advisable to provide them with access to harm reduction measures for risky sexual behavior. Given the difficulty of quitting recreational pregabalin use and the low adherence of substance users to the existing mental health care system, it is recommended to improve this system and introduce harm reduction programs.

## INTRODUCTION

Substance use disorders lead to serious illnesses, public health problems and social problems [1]. According to a report by the United Nations Office on Drugs and Crime, the number of people using psychoactive substances increased by 20% over a ten-year period, reaching 292 million in 2022. Of these, 22% have substance use disorders, and only 1 in 11 are receiving treatment for these disorders [2]. However, substance use increases the global burden of disease not only through substance use disorders, but also through an increased risk of other diseases and accidents. Global DALYs associated with substance use are also high for injuries, cardiovascular diseases, cancer, cirrhosis and other chronic liver diseases, HIV infection and tuberculosis [3]. Strengthening measures to prevent and treat substance use disorders is one of the Sustainable Development Goals [4].

Pregabalin is a gabapentinoid approved by the FDA in 2004 for the treatment of a fairly limited range of diseases, but the drug has achieved significant commercial success and entered the top 10 global pharmaceutical sales in 2017. However, this commercial success has not been accompanied by safe, prudent, and cost-effective use [5]. Reports of pregabalin’s addictive potential began to appear immediately after the drug’s market launch in 2005 [6], the first cases of pregabalin abuse in Europe were recorded in 2008 [7], and the first pregabalin use disorders were diagnosed in 2010 [8].

Acute pregabalin intoxication can lead to several complications, including liver damage [9], a range of neurological and psychiatric disorders [10, 11, 12, 13, 14, 15], respiratory depression [16], coma and death [17], and the number of emergency medical calls due to pregabalin use is increasing every year [18]. Chronic abuse of pregabalin leads to substance dependence [19], damage to the central nervous system [20], reproductive disorders [21], and significantly increases cardiovascular risk [22]. Pregabalin also has a teratogenic effect [23].

After discontinuation, withdrawal syndrome develops, including psychopathological, vegetative, neurological, and other physical symptoms. Withdrawal syndrome occurs in people of different ages and genders, with and without a history of mental disorders and substance abuse. There have been reports of withdrawal syndrome occurring even after discontinuing two weeks of pregabalin treatment at therapeutic doses [24].

In addition, pregabalin has several effects on sexual behaviour, making it a potential agent for chemsex. Chemsex is associated with an increased risk of transmission of HIV, viral hepatitis and other sexually transmitted infections [25], as well as the development of substance use disorders [26], other mental disorders [25], cardiovascular disease [26], negative social consequences [27], physical and sexual violence [28].

Given the above, recreational use of pregabalin poses a significant public health problem and exacerbates the burden of infectious and non-infectious diseases. Governments, researchers and non-governmental organisations around the world are concerned about the problem of recreational use of pregabalin and chemsex, noting the growing prevalence of these phenomena and the need to develop effective interventions for prevention and harm reduction.

The aim of this study is to examine patterns of recreational use of pregabalin in Ukraine to develop recommendations and identify interventions necessary to prevent such use and harm reduction.

The objectives of the study are:

1. To review the literature on recreational use of pregabalin, its negative consequences, and global interventions to reduce the harm caused by such use, as well as the prevention and treatment of mental and physical health disorders resulting from pregabalin use.
2. To determine the popularity of pregabalin in the information space and the awareness of pregabalin users about the dangers of recreational use of pregabalin.
3. To identify sources and assess the availability of pregabalin for recreational use in Ukraine.
4. Identify patterns of recreational use of pregabalin in Ukraine and assess the risk of negative consequences for individuals and society as a whole as a result of such use.
5. Investigate the possibility of using pregabalin as a tool for chemsex and the use of harm reduction measures for risky sexual behaviour during sexual contact under the influence of pregabalin.
6. Identify the needs of pregabalin users and assess the level of trust in society and existing institutions in Ukraine in case of need for assistance in quitting recreational use of pregabalin.

The object of the study is the phenomenon of recreational use of pregabalin in Ukraine.

The subject of the study is the impact of recreational use of pregabalin on the physical and mental health and socio-economic sphere of people who take it for recreational purposes.

Data collection was carried out using semi-structured interviews with people who use pregabalin recreationally or who have a history of such use. Based on the scientific data reflected in the «Literature Review» section of this work, a data collection tool was created. The collected data was analysed using a phenomenological method, which allowed for a deep dive into the subjective experience of each study participant and provided comprehensive answers to the questions necessary to complete the study tasks.

## LITERATURE REVIEW

Pregabalin is an antiepileptic drug used to treat neuropathic pain, partial epileptic seizures, fibromyalgia, and generalised anxiety disorder [29]. Structurally, pregabalin is an analogue of gamma-aminobutyric acid, but it has not been shown to bind to any subtype of GABA receptors or other 38 receptors and ion channels common in the central nervous system [30]. The mechanism of action of pregabalin is to bind to the α_2_δ subunits of voltage-gated calcium channels in the central nervous system, which reduces the depolarisation-associated flow of Ca^2+^ ions into presynaptic neurons and, as a result, the release of excitatory neurotransmitters [31]. However, researchers and the manufacturer itself have stated that this is not the only mechanism of action, and the exact mechanism of action remains unknown [32, 33].

Despite its limited range of FDA-approved indications, the substance has been widely used in medicine, predominantly off-label [34, 35, 36, 37, 38, 39, 40, 41, 42, 43], facilitated by the manufacturer’s illegal promotion of the drug [44]. The FBI proved that the manufacturer had engaged in aggressive marketing and paid kickbacks to doctors for prescribing a number of drugs, including pregabalin. The case was called the most high-profile healthcare fraud case ever, and the fine imposed was the largest in US history [45].

Pregabalin ranked tenth in global sales and Pfizer became the largest pharmaceutical company in terms of total sales in the 2017 [46]. Analysts predict that sales of pregabalin will continue to grow steadily [47, 48]. In Ukraine, as of March 2025, pregabalin-containing drugs were included in the list of the most commonly used medicines [49].

Among the clinical and side effects of pregabalin, there are a number that form the basis for its recreational use, in particular its anti-anxiety and hypnotic effects [50, 51], dissociative effects and euphoria [7], psychomotor stimulation [51, 52], increased sociability [53], sexual disinhibition and increased libido [54]. There have also been reports of visual hallucinations [55] and lucid dreaming [56] under the influence of pregabalin, which may be of interest to psychonauts [57].

In addition, pregabalin has found widespread use among users of other psychoactive substances [58], either as an alternative to them or as a means of potentiating their effects [51], as well as one that can eliminate the symptoms of nicotine withdrawal [59], alcohol [60, 61, 62, 63, 64, 65], cannabinoids [66], benzodiazepines [67, 68, 69], Z-drugs [70] and opioids [58, 71, 72, 73, 74].

Reports of pregabalin’s addictive potential appeared immediately after the drug was launched on the market in 2005 [6], In 2006-2008, the first cases of abuse and the first fatal case with pregabalin detected in the blood were reported in Europe [7, 75], and in 2010, the first pregabalin use disorders were diagnosed [8]. The prevalence of pregabalin abuse subsequently began to grow rapidly in all age groups in countries around the world, particularly in Denmark [76], Spain [7], Germany [58, 77, 78, 79, 80], Portugal [81], Serbia [82], France [51, 83, 84], Sweden [85] and other European Union countries [86, 87], Australia [18, 88, 89], Algeria [90], Great Britain [75, 91, 92, 93], Israel [94, 95], Jordan [96], New Zealand [97], Saudi Arabia [98, 99], Sudan [100], the United States [101, 102, 103], Turkey [17, 104], Switzerland [105] and Japan [106]. Moreover, France has recorded a high prevalence of recreational use of pregabalin among children and adolescents aged 10–17 [107], in the United States, between 2018 and 2022, there was an increase in the number of gabapentinoid prescriptions among children and adolescents [108], and in Spain in 2022, the DDD (defined daily dose per 1,000 population) for pregabalin exceeded that for metamizole [7]. There have been reports of injecting pregabalin in Europe [109].

Although pregabalin is a prescription drug available from pharmacies, there are reports in the literature of numerous requests for it to be dispensed without a prescription or with a forged prescription [7, 90]. In Ukraine, there are no reports of non-prescription requests for pregabalin in the scientific literature, but such reports appear in news feeds and journalistic investigations [110], as well as in court practice [111]. At the same time, in France, about half of those who abuse pregabalin purchase it with a genuine prescription, which confirms the growing availability of the substance. There are also reports of the substance being available on the black market [51].

Recreational use of pregabalin has been linked to numerous cases of acute poisoning, both in combination with other substances and without them. Such cases of poisoning have been reported in various countries around the world and often lead to coma and death, with their prevalence increasing every year [17, 92, 112, 113, 114, 115, 116, 117, 118, 119]. Cases of acute liver damage [9, 120, 121, 122], psychosis [11, 123, 124], encephalopathy [14], hypoglycaemic coma [125, 126, 127], rhabdomyolysis [128, 129], the development of motor disorders [10, 130, 131, 132, 133], self-harm [13, 134], suicidal thoughts and behaviour [12, 135], bullous pemphigoid [136], and status epilepticus [15] in acute pregabalin poisoning.

In Australia, the number of emergency medical calls related to poisoning associated with pregabalin use increased more than tenfold between 2012 and 2017 [18].

A correlation has been established between gabapentinoid use and the onset of mental disorders. Between 2004 and 2023, gabapentinoid use increased the incidence of mental disorders in the United States, particularly anxiety and suicidal thoughts [137]. Additional gabapentinoid therapy with the aim of prescribing fewer opioids has, conversely, increased overall opioid consumption [138].

Experimental studies in rats have demonstrated the ability of pregabalin to cause oxidative stress in the central nervous system, leading to neuron degeneration [139]. It has also been shown that the neurotoxic effect of pregabalin is similar to that of tramadol, which manifests itself in changes in the expression of dopamine receptor genes (decreased expression of the first (D1Rs) and fifth (D5Rs) types and increased expression of the second (D2Rs) and fourth (D4Rs) types), neuroinflammatory processes, degeneration of pyramidal and granular cells, and the appearance of amyloid plaques. The above mechanisms lead to brain damage, memory impairment, decreased locomotor activity, and the onset of psychosis. These effects are observed with prolonged administration of high doses of pregabalin, which is typical for recreational use [20, 140]. A decrease in cognitive function has also been shown with prolonged use of pregabalin at therapeutic doses [141].

The effect of pregabalin abuse on reproductive function has been demonstrated. In male rats, high doses of pregabalin reduce total testosterone levels, increase luteinising and follicle-stimulating hormone levels, and cause spermatogenesis suppression, morphological changes in spermatozoa, and degeneration of the seminiferous tubules. In females under the same conditions, a decrease in pituitary hormone levels and an increase in the number of atretic ovarian follicles were observed. The mechanism of toxicity is associated with the activation of oxidative stress and the subsequent development of diffuse gonadal atrophy [21, 142, 143]. Female rats mated with males receiving pregabalin showed a significant decrease in placental weight, as well as a decrease in foetal weight, length and viability [21]. A number of experimental studies using test animals have shown that pregabalin causes inhibition of foetal limb differentiation, leading to congenital abnormalities of the limbs, spine, craniofacial abnormalities, and teeth [144, 145, 146]. Pregabalin also affects the neurogenesis and morphogenesis of ventral dopaminergic neurons in the midbrain of the foetus [147]. Pregabalin has also been shown to have a teratogenic effect in rats, with the development of degenerative changes in the heart, blood vessels, kidneys, and organs of vision [148, 149].

A link has been established between prenatal exposure to pregabalin and an increased risk of congenital anomalies and delayed neurological development in humans, but these findings require further investigation [23].

Long-term use of pregabalin increases cardiovascular risk, particularly the risk of deep vein thrombosis, peripheral vascular disease, pulmonary embolism [22], acute heart failure, and decompensation of chronic heart failure [150, 151, 152], atrioventricular block [153, 154]. In rats, it has been proven that pregabalin leads to pronounced oedema and vacuolar changes in cardiomyocytes [155].

Taking pregabalin is associated with an increased risk of falls [36], bone fractures [156] injuries, and traffic accidents [157].

After discontinuing pregabalin, withdrawal syndrome develops, including psychopathological (insomnia, dysphoria, delusions, visual and auditory hallucinations, thoughts of self-harm, anxiety, suicidal thoughts, feelings of insanity, asthenia, agitation, alexia, distorted colour perception, akathisia, hypotimia, anhedonia, impulsivity, aggressiveness), vegetative (tremor, shortness of breath, palpitations, dizziness, sweating, arterial hypertension, tachycardia, lacrimation), neurological and other physical symptoms (chest pain, weakness and pain in the legs, tonic-clonic seizures, headache, coordination disorders, nausea, lack of appetite, discomfort in the epigastric region, suffocation, chills, physical discomfort). There have been reports of withdrawal syndrome occurring after discontinuation of two weeks of pregabalin treatment at therapeutic doses. Withdrawal syndrome occurs in people of any age, gender, with or without a history of psychiatric disorders or substance use [19, 24, 158]. In addition, pregabalin may be a potential agent for chemsex. The term “chemsex” is defined as the voluntary use of psychoactive and non-psychoactive substances in a recreational context before or during sexual intercourse to facilitate, prolong, or enhance the sexual experience [27, 159]. Today, chemsex is a growing public health concern worldwide [26, 160], and in France, a high prevalence of chemsex has been reported among young students [161]. Chemsex is associated with an increase in risky sexual behaviour [25], more frequent casual sexual encounters [27], unprotected sex with multiple partners [162], open relationships, group sex, double penetration, fisting [163, 164], transactional sex, sharing sex toys [164], unprotected sex [165, 166, 167], including anal sex [168], higher consumption of alcohol and injecting drugs [164]. Accordingly, chemsex is associated with an increased risk of transmission of HIV, viral hepatitis, and other sexually transmitted infections [25, 164, 167, 169, 170, 171, 172, 173], as well as the emergence of disorders related to psychoactive substance use [26, 170], other mental disorders [25, 27], cardiovascular disease [26], and negative social consequences [27]. In addition, prolonged sexual contact under the influence of certain substances increases the risk of rectal, penile, and vaginal injuries, which in turn increases the risk of sexually transmitted infections [159, 174]. It has been shown that people who engage in chemsex have a significantly higher risk of experiencing physical and sexual violence [28].

Research using semi-structured and in-depth interviews to collect data has found that substances used for chemsex are intended to enhance sexual desire and subjective arousal, increase the perception of intimacy and satisfaction from the sexual experience, prolong sexual intercourse, lead to sexual disinhibition, reduce the fear of rejection by a potential partner, and serve as a means of overcoming society’s rejection of such behaviour [175, 176]. Pregabalin meets all of these criteria, as it disinhibits the sexual sphere and increases libido [19, 54], increases sociability [53], has anti-anxiety [50] and psychostimulant effects [51, 52], and leads to delayed ejaculation and anorgasmia [177, 178, 179]. However, no references to the use of pregabalin as a tool for chemsex could be found in the scientific literature.

Governments and researchers around the world are concerned about the recreational use of pregabalin, noting its growing prevalence and consumers’ often unsuccessful attempts to stop taking the drug.

Doctors suggest various treatment strategies for disorders associated with pregabalin use in psychiatric hospitals using drugs from other groups, report successful cases, and emphasise the importance of early intervention [19]. However, such treatment can take up to several months [180, 181, 182], which increases the risk of relapse and places an additional burden on the healthcare system.

The issue of chemsex has also not gone unnoticed by countries around the world. There are an increasing number of reports in scientific literature on the need to address this issue and develop effective harm reduction interventions [170, 183, 184, 185]. In Belgium, the Chemfield project was launched to develop a digital support tool to reduce the harm caused by chemsex practices [186], and in Taiwan, a medical centre dedicated exclusively to chemsex issues was established in 2017 [187].

The trafficking of pregabalin is restricted in Argentina, the United Kingdom, Armenia, Jordan, Norway, the United Arab Emirates, Saudi Arabia, the United States of America, the Russian Federation, Turkey, France, and Sweden [51, 188], but despite this, the prevalence of gabapentinoid use is growing rapidly [17]. In Ukraine, although pregabalin is available by prescription in accordance with the law, it is not included in the list of narcotic drugs, psychotropic substances and precursors approved by Resolution of the Cabinet of Ministers of Ukraine No. 770 of 6 May 2000 “On the Approval of the List of Narcotic Drugs, Psychotropic Substances and Precursors” and is not included in any of the tables of narcotic drugs, psychotropic substances and precursors in illegal circulation, approved by Order of the Ministry of Health of Ukraine No. 188 of 1 August 2000 “On the Approval of Tables of Small, Large and Especially Large Quantities of Narcotic Drugs, Psychotropic Substances and Precursors in Illegal Circulation”. Moreover, according to Decree of the President of Ukraine No. 82/2025 “On the decision of the National Security and Defence Council of Ukraine dated 12 February 2025 “On additional measures to ensure the availability of medicines for Ukrainians”, additional measures were implemented to increase the availability of a number of medicines, including those containing pregabalin [49]. In Australia, the United Kingdom and Canada, the reduction in the price of pregabalin as a result of subsidies and the emergence of cheaper generic drugs containing pregabalin has led to an increase in the prescription of pregabalin [36].

Given the above, recreational use of pregabalin poses a significant public health problem, exacerbates the burden of infectious and non-infectious diseases, and requires further study and development of effective interventions aimed at preventing recreational use of pregabalin and reducing the harm caused by such use.

## METHODOLOGY

Since recreational use of pregabalin, like any recreational use of psychoactive substances, is a complex social, psychological and behavioural phenomenon, and no scientific data on recreational use of pregabalin specifically in Ukraine could be found, qualitative methods were chosen for this study. This will allow for a deeper understanding of the problem and reveal hidden aspects that may be overlooked when using quantitative methods.

The criterion for inclusion in the study is recreational use of pregabalin at the time of the study or episodes of such use in the past.

Since the issue of psychoactive substance use in Ukraine is stigmatised, it is difficult to access the target group. Therefore, the sample was formed using the “snowball” method, where the initial respondents were asked to provide recommendations and contacts of other persons who met the criteria for inclusion in the study.

Data collection was carried out using semi-structured interviews with a guide developed on the basis of existing scientific literature. Data collection was carried out until saturation point was reached. Semi-structured interviews were conducted in a separate room without access to third parties, alone with the respondents. Respondents were assigned identification numbers, and all data collected was linked only to these numbers; personal data was not included in the data collection forms. Such interviews were conducted with ten respondents who met the criteria for inclusion in the study.

The guide includes six sections of questions that enable answers to be provided to the questions set out in the research objectives:

– introduction;
– history of use. This section aims to identify possible risk factors for the onset of recreational use of psychoactive substances, and pregabalin in particular, to assess consumers’ awareness of the dangers of such use, and to study patterns of recreational pregabalin use and the risks of negative consequences for individuals and society as a whole;
– availability of pregabalin. The questions in this section will allow us to investigate the availability and sources of pregabalin for recreational use in Ukraine;
– pregabalin and chemsex. This section will allow us to answer the question of whether pregabalin has the necessary effects for its use in chemsex and to find out whether pregabalin is used for chemsex in Ukraine;
– consequences of use. This section will answer questions about the negative consequences of recreational use of pregabalin for health, social and other areas of human life. This section also includes questions about quitting recreational use of pregabalin, the onset of withdrawal symptoms and the success of users in quitting such use;
– needs and necessary assistance. This section will answer questions about the need for assistance in quitting recreational use of pregabalin and the possibility of obtaining such assistance in Ukraine, assess the level of trust of pregabalin users who are trying to quit recreational use in society and the authorities available in Ukraine.

The data was analysed using a phenomenological method, which allowed us to delve deeper into the subjective experiences of respondents and identify universal structures in these experiences in order to understand the interventions that need to be directed towards solving the problem of recreational pregabalin use in Ukraine.

The subject of the study is quite sensitive, since recreational use of psychoactive substances in Ukraine is stigmatised and, in some respects, criminalised, and the consequences of such use can be emotionally negative and traumatic for respondents. The study has a number of risks and limitations, in particular:

– difficulty in accessing respondents;

– incomplete and unreliable data;

– psychological consequences for respondents;

– legal aspects related to the circulation of psychoactive substances.

The chosen methodology also creates a number of risks and limitations, in particular:

– subjectivity of the sample, as respondents will recommend people from their social circle, which may lead to the involvement of a certain homogeneous group;

– risk of interruption of the flow of respondents;

– subjectivity of interpretation and bias of the researcher.

The research protocol was reviewed by the Research Ethics Committee of the National University of Kyiv-Mohyla Academy and was approved by Protocol No. 6 of 18 December 2024.

## RESULTS

### Demographic characteristics

Of the 10 respondents, 6 were male and 4 were female. The average age was 24 (min=18, max=32). The level of education ranged from basic general secondary education (9 years of school) to a master’s degree.

None of the respondents were employed. This information is reflected in more detail in Table 1.

**Table 1.** Demographic characteristics of respondents.

*Demographic characteristics of respondents*
| ID | Sex | Age range | Education | Employment |
| --- | --- | --- | --- | --- |
| 1 | M | 23-27 | Post-secondary non-tertiary | Unemployed |
| 2 | M | 18-22 | Complete general secondary education | Bachelor student |
| 3 | M | 23-27 | Master | Unemployed |
| 4 | M | 23-27 | Bachelor | Unemployed |
| 5 | M | 23-27 | Bachelor | Unemployed |
| 6 | F | 18-22 | Post-secondary non-tertiary | Bachelor student |
| 7 | F | 18-22 | Lower secondary | Vocational student |
| 8 | M | 33-37 | Bachelor | War veteran |
| 9 | F | 23-27 | Bachelor | Unemployed |
| 10 | F | 18-22 | Lower secondary | Vocational student |

Nine respondents reported having psychopathological symptoms prior to starting pregabalin use, four of whom had sought emergency and planned psychiatric care due to psychoactive substance use, and three had sought psychological help.

After starting pregabalin, one respondent had sought emergency medical care for acute psychotic symptoms while using pregabalin with other psychoactive substances, and three respondents had sought scheduled psychiatric care while using pregabalin with diagnosed: depressive episode (one respondent), borderline personality disorder (two respondents) and obsessive-compulsive disorder (one respondent). All respondents have a history of psychoactive substance use.

All respondents began using psychoactive substances with alcohol, with a mean age of onset of alcohol use of 13 years (min=9, max=16). The average age at which respondents began using psychoactive substances (other than alcohol and tobacco) was 14 years (min=11, max=17), and pregabalin was 21 years (min=14, max=29). A number of respondents reported systematic use of certain psychoactive substances, including eight respondents who reported systematic use of marijuana, five who reported systematic use of amphetamine, two who reported systematic use of mephedrone (one of the respondents used it intravenously), two reported regular use of benzodiazepines, one reported regular use of cocaine, one reported regular use of ecstasy, one reported regular use of kratom, one reported regular use of methadone by injection, and one reported regular use of LSD before trying pregabalin. One respondent reported that pregabalin was the first psychoactive substance (other than alcohol and tobacco) she had used in her life (being a minor). A more detailed profile of respondents’ recreational use of psychoactive substances is provided in Figure 1, with more detailed information on respondents in Appendix A. Psychoactive substances are classified according to the British “Drugs Wheel” model, which is based on the psychoactive effects, mechanisms of action and harm reduction measures for each group of psychoactive substances [189].

**Figure 1.**
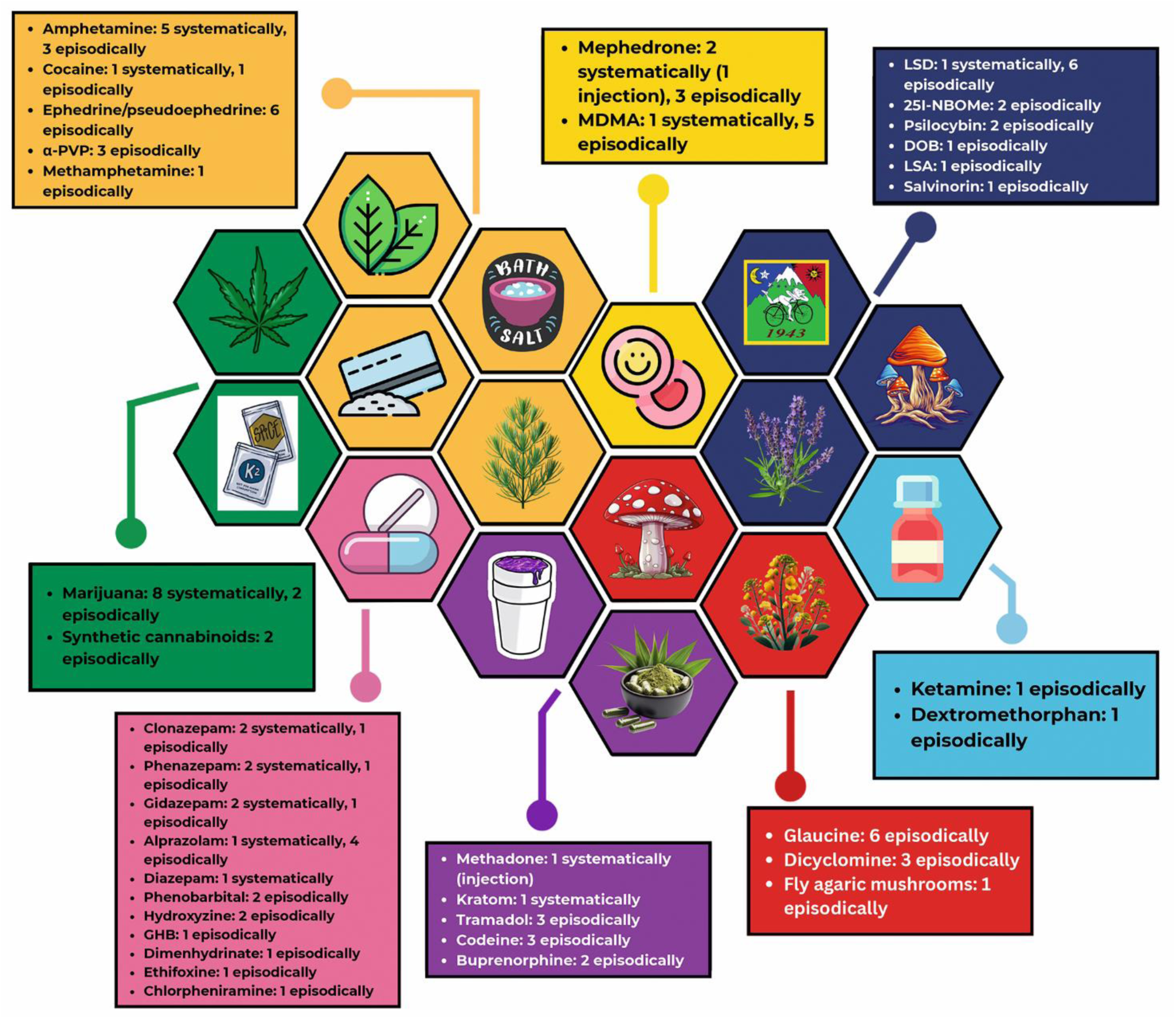
Profile of recreational use of psychoactive substances by respondents.

**Figure 2.**
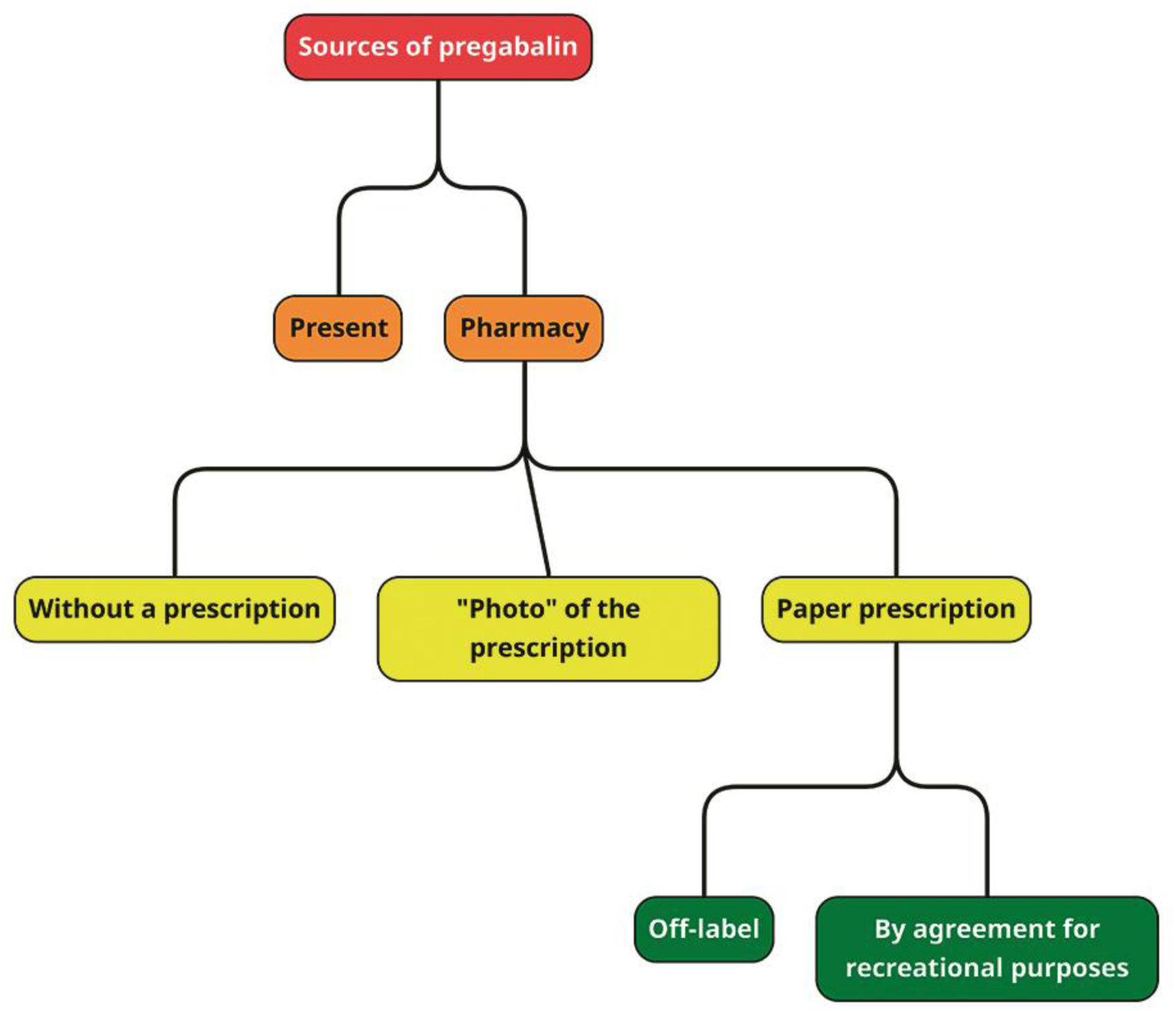
Sources of pregabalin for recreational use.

### Pregabalin in the information space

Eight respondents learned about pregabalin from acquaintances who used it regularly, one respondent from a piece of music, and one from social media. Most respondents report the popularity of recreational use of pregabalin among their peers and the growing popularity of pregabalin in society:

*«It must be an epidemic after all. Because everyone I’ve ever used psychoactive substances with has either tried pregabalin, used it for quite a long time, or still uses it to this day» - respondent No. 1, male, age range 23-27 years old, Vinnytsia*

*«At that time (2024), everyone around me was either regularly using pregabalin or had at least tried it» - respondent No. 3, male, age range 23-27 years old, Vinnytsia*

*«Pregabalin, just in the last six months, I get the impression that there’s been some kind of boom in society» - respondent No. 4, male, age range 23-27 years old, Vinnytsia*

*«It’s really very popular. When I was 15, I was surprised that such a substance existed, but then I was even more surprised because I met people, we started talking, and it turned out that they also used it. Very often. Especially among teenagers» - respondent No. 7, female, age range 18-22 years old, Vinnytsia*

*«This drug has recently become very widespread in society as a whole, with many people using it regardless of age. These include people over 50, many military personnel after hospitalisation, and many of those currently serving. Many civilians, teenagers, and girls also abuse it» - respondent No. 8, male, age range 33-37 years old, Vinnytsia*

*«For doctors, it’s like medicine No. 1 for everything. Many of my friends started taking it as prescribed by their doctor and didn’t know at the time that it (pregabalin) could be used recreationally. But then they found out about it and started abusing it. I don’t have any friends who have never tried pregabalin» - respondent No. 9, female, age range 23-27 years old, Vinnytsia*

*«It (pregabalin) is very common, especially among teenagers, because teenagers do not yet know where to get certain substances, but there are pharmacies where you can easily buy something» - respondent No. 10, female, age range 18-22 years old, Vinnytsia*

In addition, respondents report an increase in the amount of entertainment content on social media devoted to the recreational use of pregabalin:

*«It (pregabalin) began to be frequently misused and promoted to the masses through TikTok and media personalities. Of course, the emphasis was more on alprazolam, but pregabalin also began to be mentioned more and more often» - respondent No. 3, male, age range 23-27 years old, Vinnytsia*

*«Also, when I started taking pregabalin, every video I saw on TikTok was about 13-year-old girls buying up the entire pharmacy» - respondent No. 7, female, age range 18-22 years old, Vinnytsia*

*«There are now a lot of videos on TikTok about pregabalin, and there are many more trip reports about pregabalin. People also often use words derived from pregabalin in their social media signatures» - respondent No. 10, female, age range 18-22 years old, Vinnytsia*

### Respondents’ awareness of pregabalin

Seven respondents knew nothing about pregabalin when they first took it and were unaware of the risks associated with its recreational use:

*«They just told me, «It’s dope» and I agreed» - respondent No. 3, male, age range 23-27 years old, Vinnytsia*

*«I didn’t know what it was before, and I didn’t know when I took it. They just said,*

*«here’s a pill, take it, it’ll be fun»» - respondent No. 4, male, age range 23-27 years old, Vinnytsia*

*«I didn’t really know. Even when I tried it, I didn’t know what I had tried» - respondent No. 5, male, age range 23-27 years old, Vinnytsia*

*«I brought up the topic of how it would be interesting to try something new during the winter holidays, like some pills. He immediately took pregabalin out of his back pocket and suggested we do it right away. I took two pills» - respondent No. 7, female, age range 18-22 years old, Vinnytsia*

*«I knew nothing at all. I had no acquaintances who could tell me about it and how it might all end» - respondent No. 9, female, age range 23-27 years old, Vinnytsia*

Three respondents were aware of the risks to some extent, but decided to try it anyway:

*«I understood that it was very bad, I understood that people become dependent on it (pregabalin) and that it is a chemical. It was scary to try it for the first time because I didn’t know what to expect» - respondent No. 2, male, age range 18-22 years old, Vinnytsia*

*«I was afraid of overdosing, but after the first use, all my fears disappeared. Before that, I was afraid of various pharmacy substances, tablets, syrups» - respondent No. 6, female, age range 18-22 years old, Vinnytsia*

*«I knew that people abuse it (pregabalin) and that there are some negative consequences, but I didn’t know what they were specifically, I wasn’t interested» - respondent No. 8, male, age range 33-37 years old, Vinnytsia*

After starting to take pregabalin, respondents had the impression that the substance was safe:

#### «Until then, I had been able to control my use of psychoactive substances, but with pregabalin, I got caught in a trap. That’s because when you take pregabalin, you

*don’t realise what might happen next. It looks so innocent, as if nothing bad could come of it» - respondent No. 4, male, age range 23-27 years old, Vinnytsia*

*«I wouldn’t call it (pregabalin) a completely narcotic substance. I don’t see anything particularly negative about it» - respondent No. 6, female, age range 18-22 years old, Vinnytsia*

*«I would even say that it (pregabalin) is a harmless substance that does not cause significant harm to the adolescent body» - respondent No. 7, female, age range 18-22 years old, Vinnytsia*

*«When it comes to amphetamine, people at least understand that it’s not good and that they need to stop, but pregabalin is a medicine. It doesn’t look as scary as amphetamine, but if you think about it, it’s also very bad» - respondent No. 9, female, age range 23-27 years old, Vinnytsia*

### Availability of pregabalin

Eight respondents obtained pregabalin for their first dose from acquaintances who used it, one respondent purchased pregabalin for their first dose at a pharmacy, and one respondent obtained pregabalin from a narcologist. All respondents have experience purchasing pregabalin without a prescription at regular city pharmacies:

*«I: How did you buy pregabalin?*

*R: At a pharmacy, a regular city pharmacy*.

*I: Did you have a prescription?*

*R: No, I bought it without a prescription*.

*I: Did you have a specific pharmacy where they knew you and sold it to you?*

*R: No, it was different pharmacies all over the city» - respondent No. 1, male, age range 23-27 years old, Vinnytsia*

*«If one pharmacy did not sell it without a prescription, I would go to the next one, and they would definitely sell it there» - respondent No. 3, male, age range 23-27 years old, Vinnytsia*

*«I asked around among my acquaintances and found a pharmacy near my home. There were two pharmacists there, one of whom simply dispensed pregabalin to me without a prescription, while the other asked for a prescription but did not require me to show it» - respondent No. 6, female, age range 18-22 years old, Vinnytsia*

*«I: Is it difficult to find pregabalin?*

*R: Much easier than even alcohol. Once I went to a shop and they wouldn’t sell me cigarettes. After that, I went to a pharmacy and bought pregabalin (without a prescription)» - respondent No. 7, female, age range 18-22 years old, Vinnytsia*

*«R: I bought it at different pharmacies; some require a prescription, some don’t*.

*I: Did it ever happen that they didn’t ask for a prescription at all?*

*R: Yes, yes, it happened, it happened many times» - respondent No. 8, male, age range 33-37 years old, Vinnytsia*

*«My friend and I started using (pregabalin) and really liked it. Then we realised that you can buy it without a prescription» - respondent No. 9, female, age range 23-27 years old, Vinnytsia*

*«We went into the pharmacy, started looking for the prescription, and they didn’t even ask for it. They just sold it and that was it» - respondent No. 10, female, age range 18-22 years old, Vinnytsia*

Later, seven respondents received photos of paper prescriptions and purchased pregabalin at pharmacies using these photos:

*«R: Well, it was a recipe, let’s say, sent from heaven into my hands. That is, it’s not an official document*.

*I: Was it on paper?*

*R: No, it was digital. I just showed the photo» - respondent No. 3, male, age range 23-27 years old, Vinnytsia*

*«During the entire time I have been using prescriptions, only once did a pharmacist point out that my prescription was written for October 2025, a date that has not yet arrived» - respondent No. 4, male, age range 23-27 years old, Vinnytsia*

*«R: Then I met a guy who bought the recipe and sent it to me*.

*I: How exactly did he buy the recipe?*

*R: On the internet. A girl created a Telegram channel where she sells recipes. It’s not difficult to fake a recipe or find someone who will buy it. There’s no problem with that» - respondent No. 7, female, age range 18-22 years old, Vinnytsia*

*«For example, in Germany, pregabalin can only be purchased from drug dealers. You cannot buy it at a pharmacy without a paper prescription. There, as in our country, you cannot simply show a photo and buy it» - respondent No. 9, female, age range 23-27 years old, Vinnytsia*

Later, three respondents were prescribed pregabalin by a psychiatrist for off-label medical purposes (two for panic attacks, one to help them stop drinking alcohol) and were given a paper prescription. One respondent noted that she also received a paper prescription from a psychiatrist, but for recreational purposes by agreement:

*«Then I remembered that I have a very good friend who is a psychiatrist. I have known him for a long time and asked him to write me a prescription. Right now, I have five prescriptions with different names, dates and dosages» - respondent No. 9, female, age range 23-27 years old, Vinnytsia*

### Patterns of recreational use of pregabalin

Seven respondents used pregabalin for the first time under the influence of alcohol. The dosage for the first intake ranged from 150 to 750 mg, and the duration of daily intake ranged from 5 to 112 days. More detailed data are presented in Table 2.

**Table 2.**
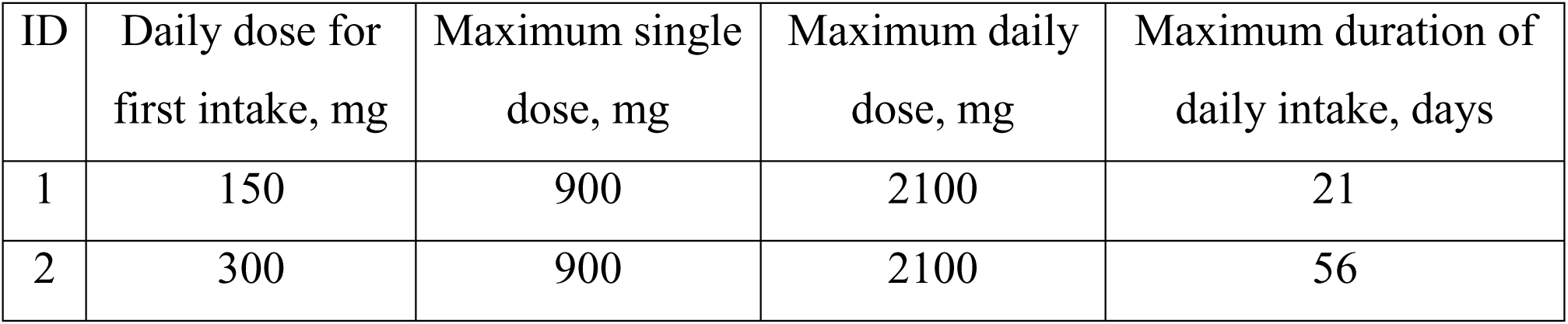
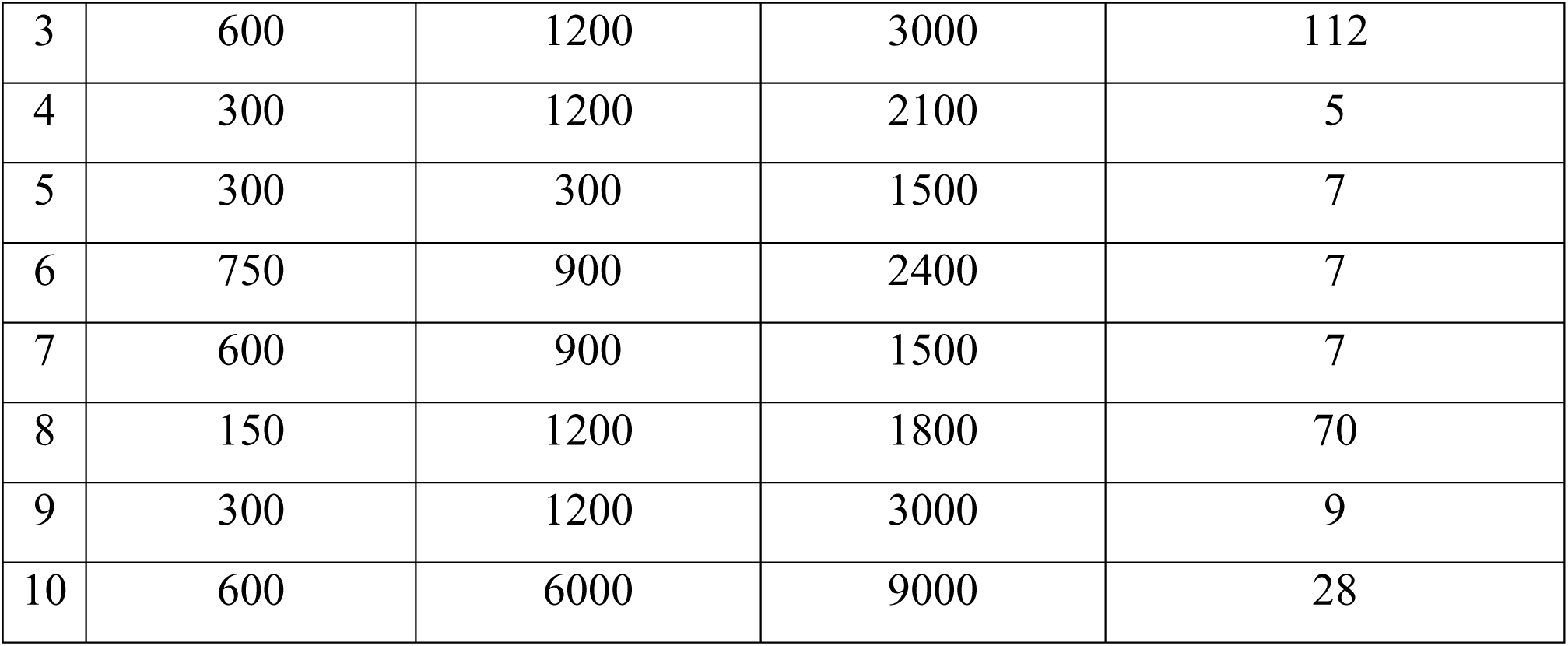
Patterns of recreational use of pregabalin by respondents.

| ID | Daily dose for first intake, mg | Maximum single dose, mg | Maximum daily dose, mg | Maximum duration of daily intake, days |
| --- | --- | --- | --- | --- |
| 1 | 150 | 900 | 2100 | 21 |
| 2 | 300 | 900 | 2100 | 56 |
| *Continuation of Table 2* |  |  |  |  |
| 3 | 600 | 1200 | 3000 | 112 |
| 4 | 300 | 1200 | 2100 | 5 |
| 5 | 300 | 300 | 1500 | 7 |
| 6 | 750 | 900 | 2400 | 7 |
| 7 | 600 | 900 | 1500 | 7 |
| 8 | 150 | 1200 | 1800 | 70 |
| 9 | 300 | 1200 | 3000 | 9 |
| 10 | 600 | 6000 | 9000 | 28 |

All respondents attempted to stop taking pregabalin, but returned to taking it after a period of time.

All respondents have experience of combining pregabalin with other psychoactive substances. Nine respondents have experience of combining pregabalin with alcohol, four respondents have experience of combining pregabalin with opioids, and two have experience of combining pregabalin with other central nervous system depressants (excluding alcohol). Six respondents have experience of combining pregabalin with marijuana, five with amphetamine-type stimulants, and five with central anticholinergics. Schematic data on the combination of pregabalin with other psychoactive substances are shown in Figure 3, with more detailed information on respondents in Appendix B.

**Figure 3.**
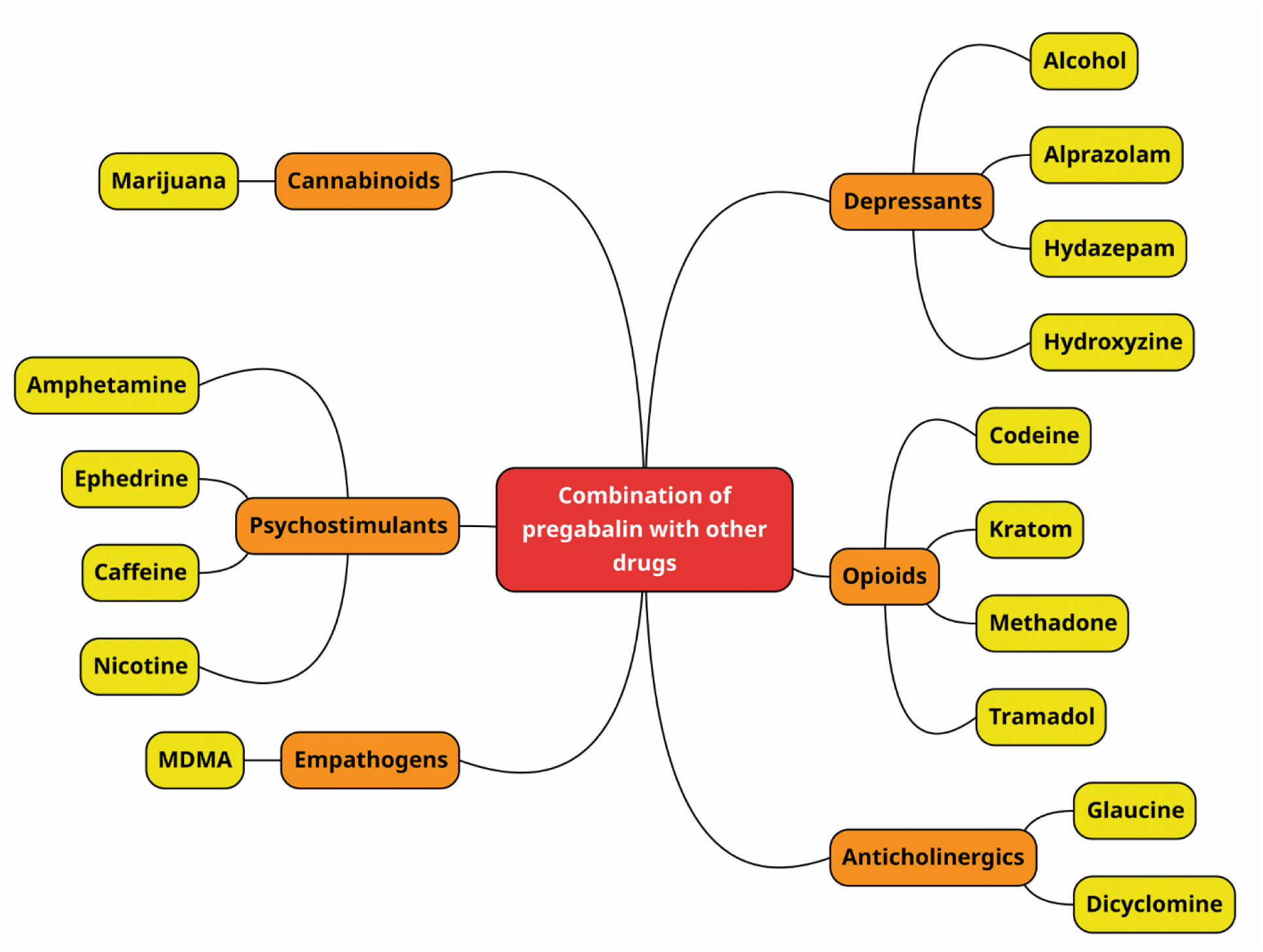
Combination of pregabalin with other psychoactive substances.

**Figure 4.**
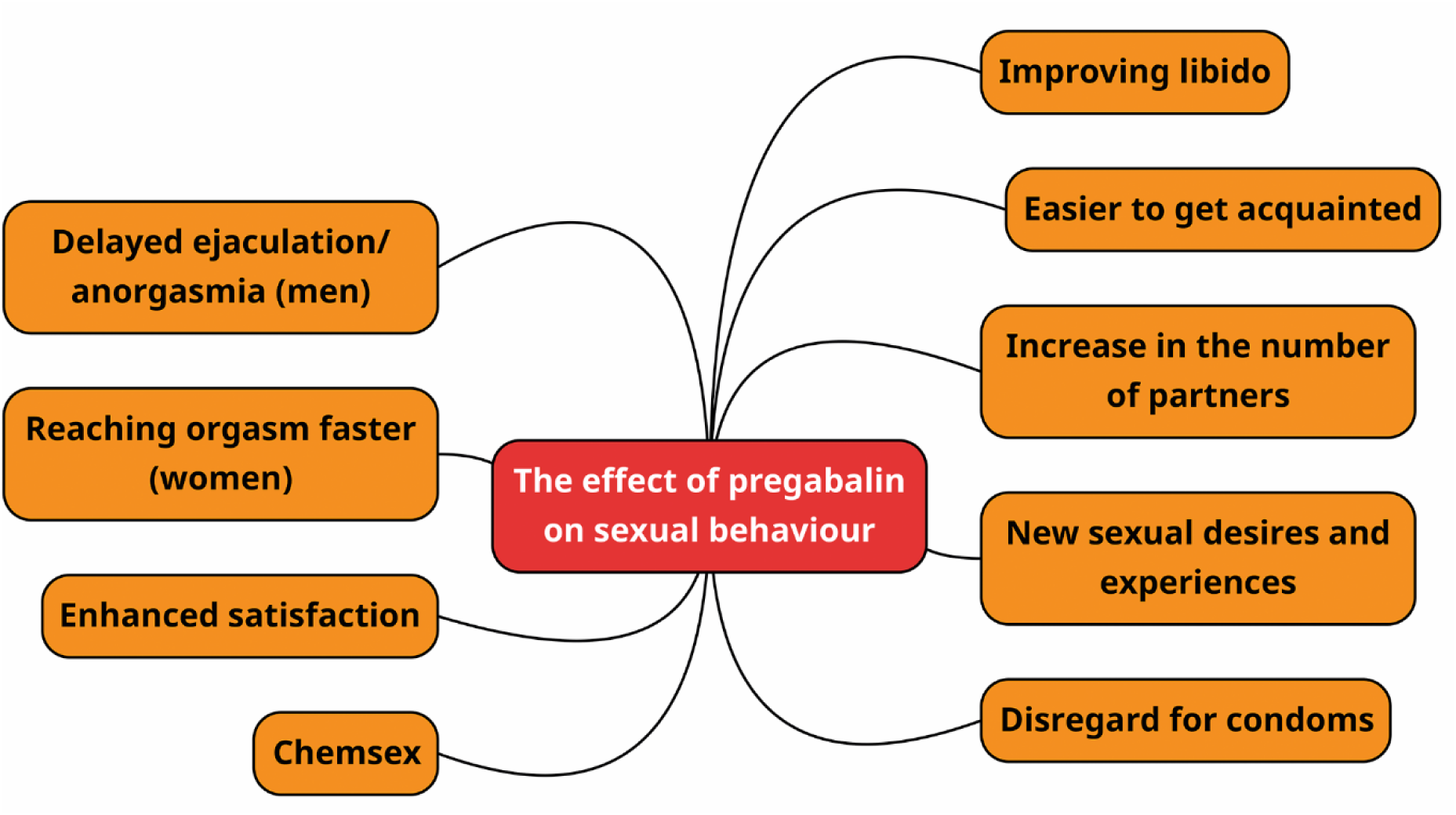
The effect of pregabalin on sexual behaviour.

Respondents also reported combining pregabalin with other psychoactive substances in order to eliminate the negative effects of using these psychoactive substances, in particular alcohol, marijuana, and dicyclomine:

*«When I took pregabalin with alcohol, I woke up very easily in the morning, although I usually feel very ill after drinking alcohol» - respondent No. 5, male, age range 23-27 years old, Vinnytsia*

*«When I use marijuana, I often start to get «paranoid» about something, thinking about things that aren’t really there. But when you mix it with pregabalin, that anxiety and «paranoia» goes away» - respondent No. 5, male, age range 23-27 years old, Vinnytsia*

*«I also mixed it with dicyclomine. Pregabalin mitigated the negative effects of dicyclomine» - respondent No. 7, female, age range 18-22 years old, Vinnytsia*

*«No hangover in the morning (after drinking alcohol) – that’s a big plus, which is why I like to mix pregabalin with alcohol» - respondent No. 9, female, age range 23-27 years old, Vinnytsia*

### The effect of pregabalin on sexual behaviour. Pregabalin and chemsex

All respondents had sexual intercourse while under the influence of pregabalin and reported subjectively positive effects on their sexual activities.

All respondents noted that with pregabalin, it is easier to meet and communicate with people, the fear of rejection by a potential partner and social disapproval of behaviour is reduced:

*«The standards disappeared, that I didn’t want to hurt the girl. Because when I’m sober, why do I need these sexual encounters if I don’t have serious intentions? And with pregabalin, I don’t care» - respondent No. 2, male, age range 18-22 years old, Vinnytsia*

*«When communicating with girls, you consider yourself more charismatic and open, without shyness or «awkwardness» - respondent No. 8, male, age range 33-37 years old, Vinnytsia*

Nine respondents reported increased sexual desire with pregabalin, two of whom reported sexual intercourse with partners they found sexually unattractive:

*«When you take pregabalin, your libido increases significantly compared to when you are sober» - respondent No. 1, male, age range 23-27 years old, Vinnytsia*

*«I: Have you had any new sexual experiences since you started taking pregabalin? R: Yes, but it was bad. There was a girl I didn’t like, but I had sex with her anyway and regretted it afterwards. We stopped talking after that» - respondent No. 2, male, age range 18-22 years old, Vinnytsia*

*«I have a girlfriend, so I’m staying away. But if I didn’t have her, it would definitely increase (the number of sexual partners), because there’s a certain “animal” attraction to other people. Stop sometimes gets lost» - respondent No. 4, male, age range 23-27 years old, Vinnytsia*

*«Once, girl and I went to a petrol station to buy alcohol. We bought the alcohol, left the petrol station, and I felt the visual effect of pregabalin that I mentioned, and along with it, my sensitivity increased significantly. I took the girl by the hand, suggested we hurry home, brought her home, and, without explaining anything, led her into the bedroom to have sex. Even though I didn’t find the girl attractive at all and didn’t want to have sex with her. But at that moment, I really wanted to» - respondent No. 5, male, age range 23-27 years old, Vinnytsia*

*«And there is another effect – you really want sex, to the point of horror. And the process itself becomes brighter, more intense, and the arousal feels 2-3 times stronger» - respondent No. 6, female, age range 18-22 years old, Vinnytsia*

*«Once, my libido increased significantly (with pregabalin) and I had sex with a girl»*

*- respondent No. 10, female, age range 18-22 years old, Vinnytsia*

Five respondents reported experiencing new sexual desires while taking pregabalin:

*«At some point, I wanted more variety. My girlfriend and I started looking for a third girl for sexual purposes. I wanted to look for some other kind of relationship with girls sexually, I became more inclined towards it» - respondent No. 1, male, age range 23-27 years old, Vinnytsia*

*«Some secret desires intensify, you want something more, sometimes you lose control» - respondent No. 4, male, age range 23-27 years old, Vinnytsia*

*«Once I suggested to my girlfriend that we try some Kamasutra positions. We weren’t usually interested in that, but with pregabalin we started looking for them and trying them out» - respondent No. 5, male, age range 23-27 years old, Vinnytsia*

*«With pregabalin, I am completely open to anything in terms of sex; I want to try something new. If the question is whether I have tried it - yes; did I like it - yes; would I have liked it without pregabalin - probably not» - respondent No. 6, female, age range 18-22 years old, Vinnytsia*

*«If you use it systematically for a long time, ordinary pleasures are no longer enough, and you want something new. It can be very scary, absurd, immoral, but at that moment, only this can provide sexual arousal» - respondent No. 8, male, age range 33-37 years old, Vinnytsia*

Five respondents reported an increased perception of intimacy and pleasure from sexual experiences with pregabalin:

*«Yes, I’ve had sex on pregabalin more than once, and it’s cool. You become more sensitive» - respondent No. 5, male, age range 23-27 years old, Vinnytsia*

*«If it does happen (sexual contact), the process itself is much more pleasant and feels more distinct. It seems to be the same thing, but the sensations are completely different» - respondent No. 6, female, 18-22 years old, Vinnytsia*

*«It feels better, it’s cool to have sex while on pregabalin. The process itself, the feeling is much more pleasant» - respondent No. 9, female, age range 23-27 years old, Vinnytsia*

Nine respondents reported an increase in the number of sexual partners after starting pregabalin, two of whom reported a sharp increase:

*«I: Has the number of your sexual partners changed since you started taking pregabalin?*

*R: It has increased. Immediately after I started taking it. After about a month, I already had three girlfriends, whereas before that I had had no sexual contact at all for two years» - respondent No. 2, male, age range 18-22 years old, Vinnytsia*

*«I: Has the number of your sexual partners changed since you started taking pregabalin?*

*R: It has increased, because I had been in a rut before that» – respondent No. 3, male, age range 23-27 years old, Vinnytsia*

Three male respondents reported an increase in the duration of sexual intercourse due to delayed ejaculation and/or anorgasmia:

*«With a higher dose, you get more stamina and it becomes more difficult to finish the sexual act. That means you can really have sex for five hours» - respondent No. 1, male, age range 23-27 years old, Vinnytsia*

*«It becomes possible not to complete sexual intercourse for several hours with pregabalin» - respondent No. 3, male, age range 23-27 years old, Vinnytsia*

*«You can’t finish for a long time, but you want to finish. Your heart is already racing and you’re thinking, how can I not die?» - respondent No. 5, male, age range 23-27 years old, Vinnytsia*

Two female respondents, on the contrary, reported a decrease in the time required to reach orgasm:

*«I know that it takes longer for men, but with pregabalin, it’s the opposite for me (orgasm is achieved faster)» - respondent No. 9, female, age range 23-27 years old, Vinnytsia*

Three respondents reported neglecting to use harm reduction measures for risky sexual behaviour while under the influence of pregabalin, although they had previously used them predominantly. One respondent always uses measures to reduce the harm from risky sexual behaviour (including under the influence of pregabalin), while the other respondents did not use them even before starting to use pregabalin.

Two respondents reported using pregabalin as a chemsex tool, namely deliberately taking pregabalin before anticipated sexual contact in order to achieve the effects of pregabalin on sexual behaviour:

*«With pregabaliт I had a pleasant time with my new girlfriend. I simply spent time with her and had sexual intercourse. Pregabalin helped me have sex with my new girlfriend» - respondent No. 1, male, age range 23-27 years old, Vinnytsia*

*«When I visit a friend, I take pregabalin not just to relax, but to improve the quality of our sex life. In everyday life, the main purpose of pregabalin is to spice up the dull routine, and in the evenings and at parties, it is usually for the purpose of better sex»*

*- respondent No. 6, female, age range 18-22 years old, Vinnytsia*

One respondent also reported intentions to use pregabalin as a tool for chemsex:

*«I did not use pregabalin specifically for this purpose, but if I stopped using it now, I might use it for this purpose at some point. However, such situations have not arisen yet, as I was always already on pregabalin» - respondent No. 3, male, age range 23- 27 years old, Vinnytsia*

### Negative consequences

Six respondents reported negative effects of pregabalin in terms of well-being after a single dose, namely nausea, vomiting, coordination disorders, spontaneous muscle twitching, numbness of the body, difficulty urinating, pain in the kidneys, liver, heart, perception, memory and attention disorders, mood swings, panic attacks, loss of consciousness:

*«There was some kind of real-time blackout. For example, you’re walking along, you see some object in the distance, but you blink and suddenly you’re crashing into it» - respondent No. 3, male, age range 23-27 years old, Vinnytsia*

*«We were travelling together in the morning, then they got off the trolleybus, I went to the back seats and lay down. It was around 10:00. Then I woke up in the same place, got off at my stop and it was already around 16:30-17:30. I don’t understand whether I lost my memory and got on and off, rode the whole time, or what, I don’t remember that moment at all» - respondent No. 3, male, age range 23-27 years old, Vinnytsia*

*«I had some kind of emotion pop up in my head, and I spent half the evening in a terrible tilt over literally nothing. I cried for the first time in two years, while under the influence of pregabalin» - respondent No. 4, male, age range 23-27 years old, Vinnytsia*

*«If you take a lot, you get a visual effect more like ecstasy, where the image starts moving back and forth. There were times when I deliberately took as much as possible to achieve this particular visual effect» - respondent No. 5, male, age range 23-27 years old, Vinnytsia*

*«I used to forget what I had just done» - respondent No. 7, female, age range 18-22 years old, Vinnytsia*

*«If the doses were high, my condition worsened. I had panic attacks, tachycardia, and generally felt unwell» - respondent No. 8, male, age range 33-37 years old, Vinnytsia*

*«There were several times when I consumed a lot at once and lost consciousness» - respondent No. 9, female, age range 23-27 years old, Vinnytsia*

*«The days pass very quickly, I don’t even notice them. Seven days pass by as if they were one. I tell people that I’ve only had a rest for two days, but they show me photos and say that a week has already passed. At first I don’t believe them, but then I look and realise that a week has indeed passed» - respondent No. 9, female, age range 23- 27 years old, Vinnytsia*

*«I lost consciousness. At that time, there was no combination, only pregabalin. I was sitting, then my cigarette fell out, and then I fell myself. I woke up the next day. I had a pulse then, but I was barely breathing» - respondent No. 10, female, age range 18- 22 years old, Vinnytsia*

When combining pregabalin with other psychoactive substances, three respondents reported the onset of acute psychotic symptoms and emergency conditions. One of them reported seeking emergency medical assistance twice for this reason:

*«When I hadn’t slept for several days and mixed substances (alcohol, pregabalin, ephedrine, glaucine), I experienced visual hallucinations. Real people appeared out of nowhere and then disappeared just as suddenly» - respondent No. 3, male, age range 23-27 years old, Vinnytsia*

*«I took 300 mg (of pregabalin), drank alcohol, smoked marijuana, lay down and couldn’t get up, lay on the street for 3-4 hours» - respondent No. 3, male, age range 23-27 years old, Vinnytsia*

*«R: Once I decided to mix absolutely everything: marijuana, amphetamine, pregabalin and alcohol. After that, I developed psychosis*.

*I: Can you describe in more detail what exactly happened?*

*R: It’s a pretty serious condition, I don’t remember much. It seemed to me that things were happening to my body that were physically impossible, for example, that my body had suddenly grown in size» - respondent No. 6, female, age range 18-22 years old, Vinnytsia*

*«It seemed to me that there were listening devices and cameras everywhere, and then hallucinations were added to that. My room changed visually, as if I wasn’t at home, and then I suddenly returned to reality. Objects in the room changed, and I didn’t understand where I was. Then I used alcohol, pregabalin, amphetamine for three days and then smoked marijuana» - respondent No. 9, female, age range 23-27 years old, Vinnytsia*

When taking pregabalin for several days or more, respondents reported the onset of thought disorders, affective disorders, suicidal thoughts and attempts, episodes of self-harm, decreased cognitive abilities, and drowsiness:

*«I gradually began to feel like I was «going mad», but I still cannot understand why these things started happening to me. It was just a stream of thoughts, I couldn’t let them go and took everything upon myself, as if I had become these thoughts. That’s why I thought I had «gone mad» - respondent No. 1, male, age range 23-27 years old, Vinnytsia*

*«Over time, the effect of «dullness» appears. You live like an amoeba, flowing from day to day, from week to week, and the further you go, the worse it gets, the less productive you are, the less desire you have to do anything, and the value of goals and tasks is lost. The whole meaning of life is lost, and you begin not to live, but to exist» - respondent No. 6, female, age range 18-22 years old, Vinnytsia*

*«Due to the fact that the dosage has to be increased, mood swings occur and the emotional background becomes less stable. The first few hours are fine, but then fear, doubts and uncertainty appear» - respondent No. 8, male, age range 33-37 years old, Vinnytsia*

*«Self-harm was still under pregabalin and alcohol. It happened before (before pregabalin) too, but not as badly. At that moment, I felt very sad, as if the whole*

*world was black, terrifying, and I didn’t like it at all. I recalled certain moments from my life, wondering why I could feel so much pain. The euphoria was gone, and I was tired of everything and everyone. I cut my legs very badly, cutting myself for about two hours. My pain threshold also increases (with pregabalin)» - respondent No. 9, female, age range 23-27 years old, Vinnytsia*

*«I was already taking pregabalin regularly at that time and decided to commit suicide. I took two blister packs at once (12600 mg), but I don’t remember much after that. It was an overdose and I ended up in intensive care» - respondent No. 10, female, age range 18-22 years old, Vinnytsia*

Respondents also report a loss of psychoactive effects after several days of daily use:

*«When I decided that I could take it on a Thursday-Saturday schedule, I felt better on Thursday. But by Saturday, when I took it, I realised that I wasn’t doing it for pleasure, but simply so that I wouldn’t feel bad. The longer you take pregabalin, the worse you feel» - respondent No. 4, male, age range 23-27 years old, Vinnytsia*

*«On the first day, everything is great because you’re clean. It stimulates you, you feel good, you’re cheerful and cool. But gradually you start taking more pills and it’s no longer enjoyable» - respondent No. 7, female, age range 18-22 years old, Vinnytsia*

*«On the first day, I feel very well. I can sleep, I can stay awake, I can rest for a whole day, drink alcohol, and feel fine. But by the sixth or seventh day, I start to feel restless and unwell, and the pregabalin no longer works as it should. That’s when conflicts start with those around me, both acquaintances and strangers, with everyone» - respondent No. 9, female, age range 23-27 years old, Vinnytsia*

Eight respondents reported negative effects after discontinuing daily use of pregabalin, although they did not report these effects before starting pregabalin, after a single episodic dose, or during systematic use of pregabalin. Negative effects included sweating, «temperature swings», chills, heaviness in the stomach, headache, vomiting, insomnia, drowsiness, tremor, weakness, fatigue, muscle twitching, constipation, perception disorders, hypothymia, anxiety, irritability, depressive symptoms, suicidal thoughts and attempts, and self-harm.

*«Pregabalin intensifies my suicidal behaviour. It has always been there, but after stopping pregabalin, I want to die much more than before I started taking it. There were times when I was already sitting on the window sill, but I always stopped myself at the last moment. I feel that if I continue taking pregabalin, at some point I may not be able to stop myself, and that is really scary» - respondent No. 9, female, age range 23-27 years old, Vinnytsia*

*«It seemed to me that the shoelace from the trainers was crawling towards me» - respondent No. 10, female, age range 18-22 years old, Vinnytsia*

More details on the profile of pregabalin use before the onset of withdrawal syndrome are given in Table 3, and a schematic representation of the manifestations of withdrawal syndrome is shown in Figure 6.

**Figure 5.**
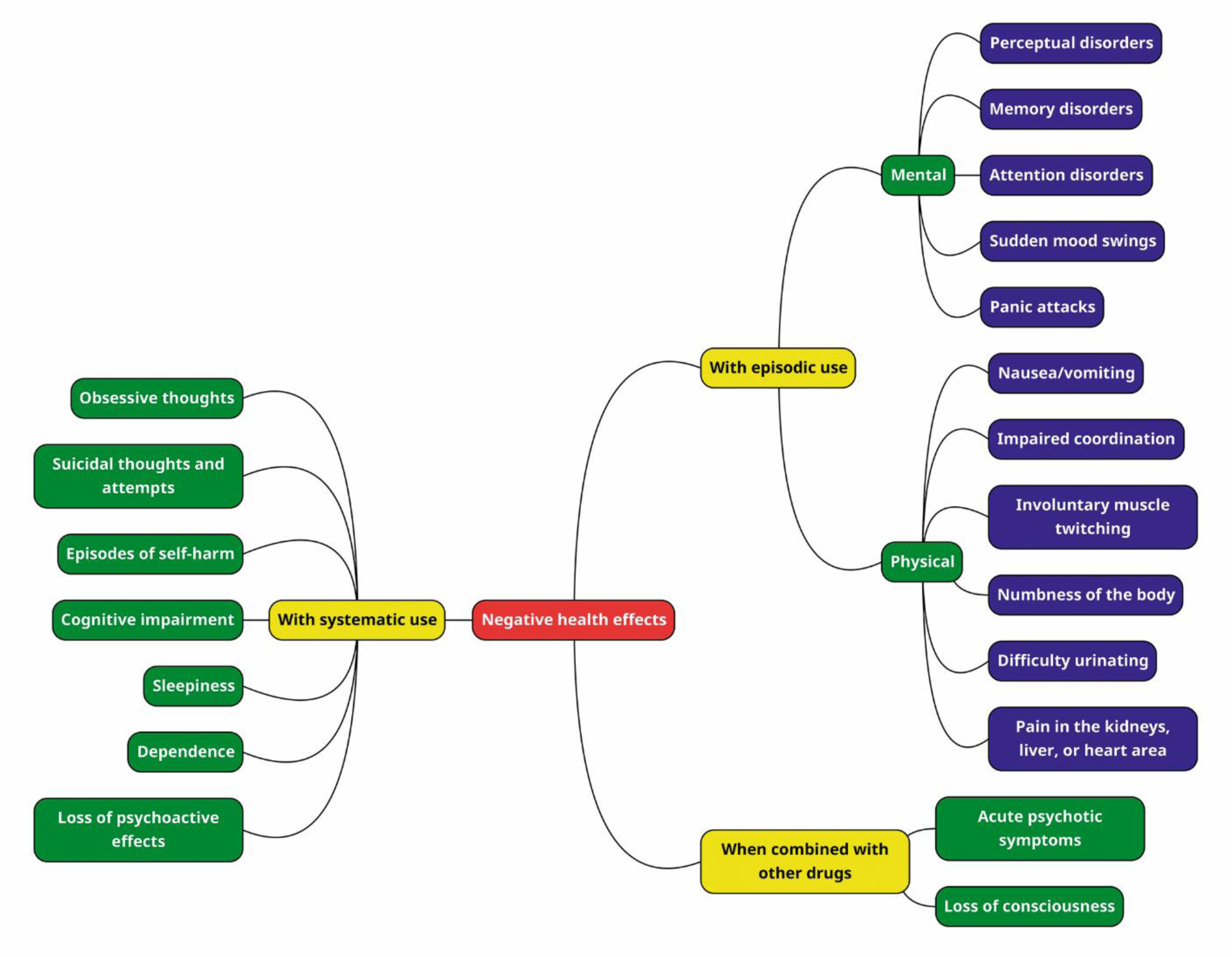
Negative consequences of recreational pregabalin use.

**Figure 6.**
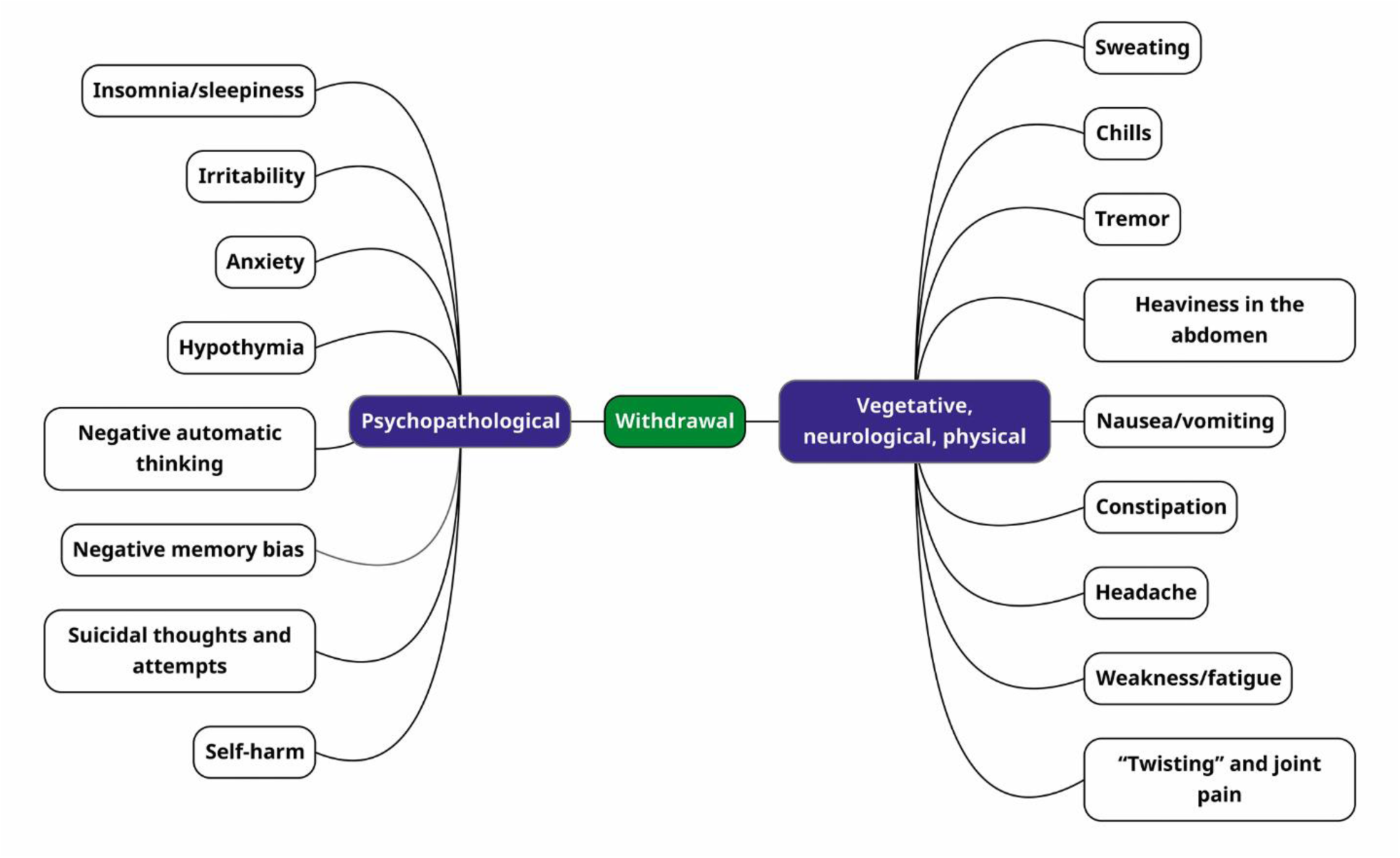
Manifestations of withdrawal after discontinuation of systematic use of pregabalin.

**Figure 7.**
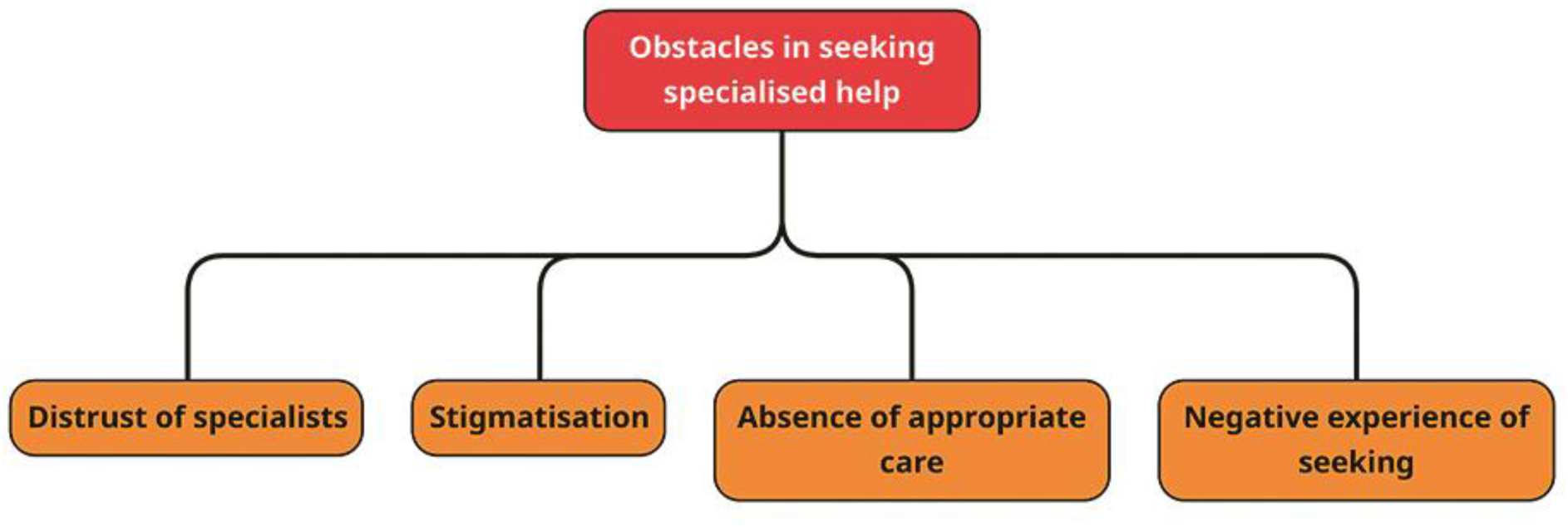
Barriers to seeking specialised help for long-term refusal to use psychoactive substances.

**Table 3.** Profile of pregabalin use before the onset of withdrawal.

| ID | Duration of daily intake, days | Daily dose on the first day of daily intake, mg | Average daily dose, mg | Daily dose on the last day of daily intake, mg | Duration of withdrawal, days |
| --- | --- | --- | --- | --- | --- |
| 1 | 21 | 300 | 900 | 600 | 7 |
| 2 | 14 | 300 | 600 | 600 | 3 |
| 3 | 56 | 300 | 600 | 600 | 14 |
| 4 | 4 | 900 | 900 | 900 | 6 |
| 7 | 7 | 300 | 600 | 300 | 1 |
| 8 | 70 | 150 | 900 | 900 | 28 |
| 9 | 4 | 750 | 750 | 750 | -* |
| 10 | 5 | 600 | 900 | 3000 | 14 |

*\* Note: Respondent No. 9 always returned to using pregabalin during withdrawal.*

Five respondents reported negative effects on their main occupation (work/study):

*«If you’re at work, everyone can tell that something is clearly wrong with you. You can see in your eyes that you’re not sober» - respondent No. 2, male, age range 18- 22 years old, Vinnytsia*

*«I lose focus very easily. I realised this when I was working while taking pregabalin*.

*In my job, the most important thing is to pay attention to what I am doing. And I noticed that I lost the desire to try. I need to have one tab open in my browser and I need to work on it. But on pregabalin, I can open 20 more tabs at the same time, and my head is filled with dreams, thoughts, etc. I look at different websites with products, and I become vague in my desires, emotions, and what I want to do» - respondent No. 4, male, age range 23-27 years old, Vinnytsia*

*«An absolute unwillingness to work (withdrawal). The work isn’t that hard, you just sit in a chair and work, but there were times when I took sick leave in the middle of a shift because I couldn’t stand it. I realised that I was just working to get through the day. But my job has very strict quality control, and I couldn’t afford to slack off» - respondent No. 4, male, age range 23-27 years old, Vinnytsia*

*«My job was directly related to communicating with people; that was my direct responsibility. Sometimes it becomes difficult (with pregabalin), it slows down my thinking a little and I don’t really feel like communicating much» - respondent No. 6, female, age range 18-22 years old, Vinnytsia*

*«I became indifferent, my productivity declined. I thought it would be better to go for a walk, shout, jump around, rather than do something useful» - respondent No. 7, female, age range 18-22 years old, Vinnytsia*

*«I lost more than one job because of pregabalin. I was dismissed from one job because they noticed I was intoxicated. Then I couldn’t go to other jobs because of withdrawal symptoms» - respondent No. 9, female, age range 23-27 years old, Vinnytsia*

*«It’s difficult to prepare my workspace; I constantly switch to something else, and everything at my workspace remains unchanged. In general, preparing my workspace (with pregabalin) takes about two hours, even though there’s nothing to prepare there at all» - respondent No. 9, female, age range 23-27 years old, Vinnytsia*

Three respondents reported some material damage in the form of loss of belongings and spending a significant amount of money (relative to their income) on pregabalin, and one respondent reported committing theft in order to obtain money to purchase pregabalin:

*«I spent a lot of money on pills. When I had an income of 1,000 UAH (about $25), all my money went to pregabalin. When I’m under the influence, I don’t count money at all. I want everything, I want to treat everyone. I could spend up to 5,000 UAH (about $120) in a day (with an income of 1,000 UAH/month). Even if it’s my last money, it doesn’t matter» - respondent No. 7, female, age range 18-22 years old, Vinnytsia*

*«It really hits the budget hard. The last time I used pregabalin for a week, I spent 22000 UAH (about $500). That’s pregabalin and alcohol. This time, I spent all the money I had: on rent, utilities, and I also exchanged euros. Now I’m in a difficult financial situation because I used pregabalin and alcohol for a week» - respondent No. 9, female, age range 23-27 years old, Vinnytsia*

*«Pregabalin became significantly more expensive, it cost a lot of money and I had to steal it. I used to steal money from my sister, I could steal someone’s wallet» - respondent No. 10, female, age range 18-22 years old, Vinnytsia*

Two respondents reported negative consequences of recreational pregabalin use on their relationships with others:

*«I don’t communicate with my family as much; I have a child. Pregabalin has had a significant impact on this; I see my child less often because I don’t want to visit them while drunk, but I can’t stop using it» - respondent No. 9, female, age range 23-27 years old, Vinnytsia*

*«My social skills are affected; I find it difficult to communicate with people and don’t want to communicate with anyone at all. I thought that other substances might be affecting this, but recently I have only been using pregabalin and alcohol. Pregabalin has definitely had an even stronger effect than anything else before» - respondent No. 9, female, age range 23-27 years old, Vinnytsia*

The respondent also reported a change in personality as a result of systematic use of pregabalin:

*«I told all my friends that they shouldn’t abuse pregabalin so much, because I noticed that it was changing them a lot. After just 1-2 months of use, they were literally different people. And then I became the same» - respondent No. 9, female, age range 23-27 years old, Vinnytsia*

### Refusal to use recreationally. Need for support

All respondents reported intentions and attempts to stop recreational use of pregabalin; three of them continue to use it regularly, while the others use it occasionally.

One respondent sought specialised help for long-term cessation of psychoactive substance use at a private rehabilitation centre, but not on her own initiative. The respondent noted the lack of specialised help at the centre and incidents of physical violence, and after returning from the centre, the respondent resumed using pregabalin:

*«I personally did not seek treatment. Once, on pregabalin, I was taken to rehab for 7 months. It is not the best place to be. There was no specialised help. Everyone who works there is a former addict who then became an employee of the centre. They have no medical or related education. They know a lot about addiction, but they don’t know how to treat it. There are no pills, no narcologist, and no doctor at all. If you get sick, you have to ask your relatives to bring you some medicine. There was violence. Once, a guy was beaten up at a weekly meeting. The staff took it calmly and said that it was part of the recovery process. According to their logic, if there is no accountability in the form of beatings, you won’t learn anything. In short, they don’t treat anything there» - respondent No. 10, female, age range 18-22 years old, Vinnytsia*

The remaining respondents did not seek specialised help for long-term abstinence from psychoactive substances. Eight respondents did not see the need to seek specialised help, while one respondent reported her intention to seek help at a private rehabilitation centre:

*«I came to the conclusion that no one but me would help me» - respondent No. 1, male, age range 23-27 years old, Vinnytsia*

*«I didn’t bring myself to such a state that I couldn’t stop on my own. I understand when it’s time to quit» - respondent No. 7, female, age range 18-22 years old, Vinnytsia*

*«I didn’t see the need. If I had already come to this decision myself (to give up psychoactive substances), then I decided that I could handle it on my own» - respondent No. 8, male, age range 33-37 years old, Vinnytsia*

*«I really want to go to a rehab now, but I don’t have the funds. I told my relatives about my addiction to pregabalin and alcohol, my episodes of self-harm, and asked for help.» - respondent No. 9, female, age range 23-27 years old, Vinnytsia*

Two of them also reported distrust of psychiatrists:

*«R: I don’t trust psychiatrists*.

*I: Trust in what sense?*

*R: I don’t think it would be effective. Most likely, the psychiatrist would prescribe some antidepressants, and I don’t think that would be effective» - respondent No. 1, male, age range 23-27 years old, Vinnytsia*

*«R: I would go to someone who doesn’t prescribe pills. I: Why don’t you want to take pills for medical reasons?*

*R: I don’t believe in them» - respondent No. 2, male, age range 18-22 years old, Vinnytsia*

Two respondents reported barriers to seeking treatment due to stigma-related reasons:

*«Once I spoke to a psychologist, she referred me to a psychotherapist, and I didn’t understand that. Did she call me abnormal?» - кespondent No. 5, male, age range 23-27 years old, Vinnytsia*

*«I cannot have any references in my documentation related to alcohol, drug or mental disorders, even basic depression, which many people suffer from. This could affect my admission to university in the future and my career» - respondent No. 6, female, age range 18-22 years old, Vinnytsia*

Two respondents reported needing help to overcome withdrawal symptoms but were unable to find appropriate specialists:

*«I had thoughts about detoxification, about getting clean, but my friend convinced me, saying that detoxification would not help» - respondent No. 1, male, age range 23-27 years old, Vinnytsia*

*«If I had known, I would have gone tomorrow without even thinking twice, to get a drip so I wouldn’t feel this withdrawal and forget about this drug» - respondent No. 4, male, age range 23-27 years old, Vinnytsia*

Two respondents reported negative experiences of their acquaintances seeking specialised help:

*«A friend of mine reached out. I think they made her even worse. She was treated in a psychiatric hospital, in a private rehabilitation centre, and took a bunch of drugs*.

*Now I see the result, and it’s awful. I don’t want to turn into something like that» - respondent No. 2, male, age range 18-22 years old, Vinnytsia*

*«Right now, I have a guy staying with me who was in a private rehabilitation centre in Kharkiv. He was there for three months, and the first thing he did after leaving was start using drugs again. He said that they beat them, bullied them, and psychologically pressured them there» - respondent No. 9, female, age range 23-27 years old, Vinnytsia*

### Other findings

Two respondents reported similarities between the psychoactive effects of pregabalin and those of opioids:

*«I was «smeared», there were physical effects, similar to tramadol» - respondent No. 1, male, age range 23-27 years old, Vinnytsia*

*«I didn’t expect any effect, but it happened and I really liked it. I compared it to opiates, that feeling of relaxation. At first, it was relaxation, and it was as if petrol had been spilled in front of my eyes, or as if it were a photo taken with some old camera. If I hadn’t known it was pregabalin, I would have thought it was some kind of codeine or other opiate. It gives you a strong feeling of relaxation, rocking you from side to side. It also relieves pain. I remember putting out a cigarette on myself and not feeling anything at all» - respondent No. 10, female, age range 18-22 years old, Vinnytsia*

The respondent also compared pregabalin withdrawal syndrome with opioid withdrawal syndrome:

*«I used it for five days: 600 mg on the first day, then 900 mg, then more than 1000 mg, and on the fifth day, I took about half a blister pack (a blister pack of 21 capsules, 300 mg each). I got tired of it and decided to stop. The next day I woke up feeling like I had a fever, my joints ached like I was on opiates. I was sweating, shivering, feeling hot and cold, my joints were twisting, I was hallucinating» - respondent No. 10, female, age range 18-22 years old, Vinnytsia*

## DISCUSSION

Pregabalin is popular among psychoactive substance users in the information space. Respondents noted that most people in their circle have a history of recreational pregabalin use. Psychoactive substance users consume pregabalin mainly to achieve euphoria, psychomotor stimulation, relaxation, anxiety relief, perceptual disturbances, chemsex, and to eliminate the negative effects of using other psychoactive substances. With the development of information technology, marketing, and targeted advertising, entertainment content dedicated to the recreational use of pregabalin and other psychotropic drugs is spreading on publicly accessible social networks, contributing to the popularity of the substance.

In addition, references to the recreational use of pregabalin are increasingly appearing in works of art, contributing to the popularity of pregabalin in artistic circles and among the target audience of consumers of such works, including adolescents. However, only the subjectively pleasant psychoactive effects of pregabalin are gaining popularity in the information space, while the risks of its recreational use are not covered. Most respondents were unaware of the risks of recreational use of pregabalin at the time of their first use, and those who had certain concerns changed their attitude towards the substance after their first use to a more frivolous one and formed the perception of pregabalin as a safe psychoactive substance. This subsequently led to a number of negative consequences, including systematic use, withdrawal symptoms, mental and physical health disorders, accidents, material damage, etc. Prescription drugs are often perceived as safer than “street” drugs, which further contributes to their abuse [190].

The combination of pregabalin with other psychoactive substances is also noteworthy. All respondents combined pregabalin with alcohol, and most respondents combined pregabalin with other central nervous system depressants and opioids. Such combinations are dangerous in terms of the occurrence of emergency conditions, overdoses and fatalities, as evidenced by scientific research data [18, 114, 191]. Most respondents also combined pregabalin with amphetamine-type stimulants, three of them noted the appearance of acute psychotic symptoms against this background, and one respondent sought emergency medical help twice for this reason. In addition, respondents reported a number of urgent conditions that arose while using both pregabalin and pregabalin in combination with other psychoactive substances, but they did not seek medical help. None of the respondents had any reservations about combining pregabalin with other psychoactive substances.

Given the low awareness among users of psychoactive substances about the risks of recreational use of pregabalin and its combination with other psychoactive substances, there is a need for educational activities among this target group. This method of prevention is recognised by researchers as one of the most recommended for reducing the prevalence of recreational use of pregabalin [100]. Scottish researchers, where a similar situation of low awareness of recreational pregabalin use has developed, also emphasise the importance of educational work on the risks of recreational pregabalin use and its combination with other psychoactive substances [192].

In addition, pregabalin has gained widespread popularity among doctors for the treatment of various nosologies and conditions, both globally and in Ukraine. Scientific data from around the world note the frequent off-label use of pregabalin [34, 35, 37, 39]. As for Ukraine, no such data could be found, but respondents reported being prescribed pregabalin for the treatment of panic attacks and alcohol abstinence, which are not indications for prescribing pregabalin, and the fact that they have a history of abuse of other psychoactive substances is a relative contraindication for such a prescription. One respondent noted that doctors often prescribe pregabalin for conditions that are not indications for its use, and that many pregabalin users in her circle of acquaintances started taking it for medical purposes as prescribed by a doctor, but then switched to recreational systematic use.

In Ukrainian medical circles, dependence on pregabalin is often considered a “myth”, with references to the possibility of unlimited safe use and the absence of interactions between pregabalin and other drugs [193]. However, a number of foreign scientific studies prove that pregabalin is addictive [8, 13, 19, 52, 80, 87, 102, 180, 181, 182, 194, 195, 196, 197, 198, 199, 200, 201, 202], and its combination with other drugs can be dangerous [36, 41, 112, 114, 115, 116, 117, 118, 192, 203, 204, 205].

It is also commonly believed that dependence on pregabalin occurs exclusively in people with a history of abuse of other psychoactive substances [206], However, the literature also describes cases of pregabalin dependence in people with no history of psychoactive substance use or psychiatric history [24]. One of the respondents also reported that pregabalin was the first psychoactive substance she tried after alcohol, even though she did not abuse alcohol and reported no psychopathological symptoms before starting to use pregabalin. After starting to use pregabalin, she went from occasional use to regular use, began trying other psychoactive substances and combining them with pregabalin.

The relevance of pregabalin’s medical use is significantly increasing against the backdrop of military operations and the growing number of patients with combat injuries. One respondent, who also has combat experience and a history of combat trauma, reported on the popularity of pregabalin abuse among military personnel and veterans after treatment and rehabilitation for combat injuries. In addition, military medics are also spreading information about the dangers of pregabalin abuse by military personnel [207].

Given doctors’ commitment to prescribing pregabalin, the lack of warnings about addiction, and the growing number of patients who could potentially be prescribed pregabalin, it is imperative to educate medical students and practising doctors about the risks of prescribing pregabalin and how to adequately assess the benefits and harms of such prescriptions. Foreign colleagues also emphasise the importance of educational work with doctors [108], but no such references have been found in the scientific literature in Ukraine.

Pregabalin is available for recreational use in Ukraine, despite prescription restrictions on the dispensing of medicines containing it. All respondents reported that pregabalin is available without a prescription in regular pharmacies in Ukrainian cities, including to minors. Respondents also reported that it is not difficult to obtain a prescription if necessary. This situation with the dispensing of pregabalin is observed in a number of countries around the world [7, 51, 90], and pharmacists report that the sale of prescription drugs without a prescription is extremely important for pharmacy profits [208].

The situation with the abuse of psychotropic drugs in Ukraine has been tense since Soviet times. Since Ukraine declared independence in 1991, the situation with the availability and abuse of psychotropic drugs has not improved; only the drugs themselves have changed.

In the 2000s, the situation with tramadol abuse gained significant publicity in Ukraine. After its appearance on the Ukrainian market, the drug was positioned as safe and non-addictive [209, 210] and was actively promoted by the medical community. Tramadol abuse was initially characteristic of consumers of other psychoactive substances as an alternative, but then more and more consumers of psychoactive substances began their path of use with tramadol [211].

The situation with recreational use of pregabalin in Ukraine today resembles the situation with tramadol at the beginning of the epidemic of its abuse, since:

1. Pregabalin is also actively promoted by the medical community as a safe drug that does not cause addiction, although scientific research data suggests otherwise.
2. Pregabalin is available for recreational use because it is sold over the counter in pharmacies, although according to the law it is available only by prescription.
3. Pregabalin is becoming the substance with which users of psychoactive substances begin their journey into substance use.

Researchers from Oxford University have also noted the similarity between the current abuse of gabapentinoids and the abuse of opioids in the 1990s, emphasising the importance of a more responsible approach to this problem in order to avoid the mistakes made in the 1990s with opioids [212].

The question of whether it is appropriate to restrict the circulation of pregabalin remains open. Researchers from around the world emphasise the need for pharmacists to comply with dispensing regulations as a method of reducing the prevalence of its recreational use [100]. In France, following the tightening of regulations on the dispensing of pregabalin from pharmacies, there has been a significant decrease in the overall dispensing of pregabalin [188], However, there is no data on the prevalence of use and the situation with pregabalin on the “black market”, while data from the United Kingdom, on the contrary, show an increase in the dispensing of pregabalin from pharmacies after the introduction of additional regulatory restrictions [36]. The experience of the “tramadol epidemic” in Ukraine has shown that the introduction of additional regulatory restrictions can lead to a shift to more dangerous patterns of psychoactive substance use. Researchers note that restrictions on the circulation of tramadol led to an increase in the prevalence of injecting synthetic opioids in Ukraine as a substitute for tramadol [213]. Thus, after tightening controls on the circulation of tramadol, other opioid receptor agonists (codeine, propoxyphene, nalbuphine), central M-cholinoblockers (tropicamide, cyclopentolate), benzodiazepines, central H1 receptor antagonists (diphenhydramine, hydroxyzine), dissociatives (dextromethorphan) and other psychotropic drugs remained available, although all of them are available by prescription according to the law. At the end of 2024, the Minister of Health of Ukraine reported on the alarming situation with nalbuphine abuse, with 3 million packages sold in 2024 and only 6,000 prescriptions issued [214], and the drug itself ranked fourth in sales in Ukraine in 2024 [215]. A similar situation occurred with tramadol in the 2000s [216].

No data on the dispensing of pregabalin in Ukraine could be found, but in 2025, drugs containing pregabalin as the active ingredient were included in the list of the 100 most commonly used drugs in Ukraine [49], which indicates that they are widely dispensed by pharmacies. As for the recreational use of pregabalin, no data on Ukraine could be found either, and the collection of such data is not included in the national monitoring of the drug and alcohol situation in Ukraine. However, the results of the study confirm the existing problem of its recreational use and access to drugs containing it in Ukraine, and the announced reduction in prices for drugs containing pregabalin only increases its availability to the population. Australia, the United Kingdom and Canada have experienced an increase in use following a reduction in the price of pregabalin as a result of subsidies and the emergence of cheaper generic drugs [36], so such a move by Ukraine could exacerbate the problem of recreational use of pregabalin.

According to scientific literature, recreational use of pregabalin leads to a number of negative consequences, which were also reported by respondents, in particular:

1. An experimental study with healthy volunteers showed a decrease in cognitive abilities when taking pregabalin for 8 weeks at a dosage of 600 mg/day [141]. Three respondents reported daily continuous use of pregabalin at a dose of 600-900 mg/day for 8, 10 and 16 weeks, respectively, confirming the existence of dangerous patterns of recreational use of pregabalin in Ukraine in terms of the risk of cognitive impairment.
2. An experimental study using laboratory animals showed the neurotoxicity of pregabalin at doses that are equivalent in humans to at least three times the therapeutic dose, i.e., at a dose of 1800 mg/day, its neurotoxic effect manifests itself [20]. One respondent reported reaching a daily dose of 1800 mg, and eight others reported exceeding this dose. These data indicate the existence of dangerous patterns of recreational pregabalin use in Ukraine in terms of the development of neurotoxic effects.
3. There have been reports of acute psychotic symptoms resulting from the use of pregabalin with other psychoactive substances [123, 124]. Three respondents also reported psychotic episodes while combining pregabalin with other psychoactive substances, and one of the respondents sought emergency medical help twice for this reason.
4. Episodes of self-harm have been described as a result of pregabalin use in people with no history of psychiatric illness or substance use [13, 134]. Two respondents also reported such episodes both during pregabalin use and after withdrawal as part of withdrawal syndrome.
5. Taking pregabalin leads to the appearance of depressive symptoms, including an increased risk of suicidal behaviour, as stated by the FDA back in 2008 [134]. A number of scientific studies prove that the use of pregabalin leads to suicidal thoughts

both during use [12, 135, 137] and after discontinuation as part of withdrawal syndrome [19, 24, 158]. Six respondents reported depressive symptoms while using pregabalin, three of whom reported suicidal thoughts, two reported suicide attempts, and one was diagnosed with a depressive episode by a psychiatrist. After discontinuing recreational use of pregabalin, six respondents reported the onset of depressive symptoms, four of them reported suicidal thoughts, and two respondents reported suicide attempts after attempting to quit recreational use of pregabalin.

1. There have been reports of the need to significantly increase the dosage of pregabalin within the first week of use in order to achieve the desired psychoactive effects [19]. Respondents also reported the need to increase the dosage during the first week of daily use due to the lack of desired psychoactive effects and increased it by 2.5-15 times during the first week, reaching a dosage of 9000 mg/day.
2. Exceeding the daily dose of pregabalin by 1000 mg is dangerous in terms of developing dependence on pregabalin [195]. All respondents reported exceeding the dose of 1000 mg/day when using pregabalin recreationally, which indicates the existence of dangerous patterns of recreational pregabalin use in Ukraine in terms of the development of dependence.
3. Numerous cases of withdrawal syndrome after discontinuation of pregabalin use have been described, with a range of psychopathological, vegetative, neurological and other physical symptoms [24]. Eight respondents reported feeling worse with similar symptoms after stopping recreational use of pregabalin. These data indicate the existence of dangerous patterns of pregabalin use in Ukraine in terms of the development of withdrawal syndrome.

One respondent’s report of committing theft in order to obtain funds to purchase pregabalin is noteworthy. Researchers around the world note that the abuse of prescription drugs leads to an increase in crime, primarily through illegal trafficking, theft and fraud, and that the recreational use of antiepileptic drugs, including pregabalin, is a particularly significant predictor of drug-related crime [190, 217]. Staff at HMP Bristol (UK) also report prisoners committing new crimes in order to obtain pregabalin [218].

The data obtained on the negative consequences of recreational use of pregabalin show that patterns of such use in Ukraine are dangerous in terms of the development of addiction, withdrawal syndrome, mental and physical health disorders, negative social consequences, accidents, and material losses. At the same time, according to the respondents, it is difficult to give up recreational use of pregabalin for a number of reasons and, accordingly, none of them had completely given up such use at the time of the study, although each had made attempts to do so, including seeking specialised help. It is also noteworthy that respondents compared the psychoactive effects of pregabalin with those of opioids, and pregabalin withdrawal syndrome with opioid withdrawal syndrome. Since the exact mechanism of action of pregabalin remains unknown [32, 33], the neurotoxic effects of pregabalin are similar to those of tramadol [20], and pregabalin has been shown to relieve opioid withdrawal syndrome [58, 71, 72, 73, 74] and induce respiratory depression [16, 219]. Withdrawal syndrome after discontinuation of pregabalin includes a physical component [24], and the phenomenon of pregabalin abuse today resembles the situation with opioid abuse at the beginning of the epidemic, both in Ukraine and worldwide [212]. There is a need for more detailed study of the pharmacodynamics of pregabalin and the pathogenesis of pregabalin use disorder in order to develop protocols and strategies for the prevention and treatment of such disorders, as well as measures to reduce the harm from recreational pregabalin use.

Some experts draw an analogy between the current situation with pregabalin and that with tianeptine before 2014 [220], which since the 1980s had been positioned as a serotonin reuptake inhibitor and used to treat depression in a number of countries around the world, until its agonism to μ-opioid receptors was proven in 2014 [221]. After that, drugs containing it were withdrawn from circulation in a number of countries, and it began to be sold as a “designer drug” in the form of a dietary supplement, mainly at petrol stations, for which it was slangily named “gas station heroin” by the FDA [222].

No data on the use of pregabalin as a chemsex tool could be found in scientific literature, but the results of the study show that pregabalin is used as a chemsex tool by both women and men. This is due to its psychoactive effects and meets the requirements that a substance must have for this purpose, namely, increasing sexual desire, subjective arousal, perception of intimacy, enjoyment of sexual experience, prolonging sexual intercourse, reducing fear of rejection by a potential partner, and overcoming society’s rejection of sexual behaviour [175, 176]. As a result, respondents reported an increase in the number of sexual partners and casual sexual encounters after starting to use pregabalin, as well as the emergence of new sexual desires and experiences. At the same time, respondents reported neglecting harm reduction measures for risky sexual behaviour under the influence of pregabalin, although they had predominantly used them before starting pregabalin use. These findings suggest that chemsex involving pregabalin increases the risk of negative consequences associated with chemsex in general, particularly the risk of transmission of HIV, viral hepatitis, and other sexually transmitted infections [25, 163, 167, 169, 170, 171, 172, 173], mental health disorders [25, 26, 27, 170] and physical health problems [26]. Thus, chemsex involving pregabalin use exacerbates the burden of both infectious and non-infectious diseases. Given this, it is advisable to engage pregabalin users in activities and programmes to counter the spread of HIV and reduce the harm from risky sexual behaviour and chemsex in general.

All respondents intended to stop recreational use of pregabalin, but none of them succeeded. One respondent sought help at a private rehabilitation centre, while the others did not seek specialised help. Among the reasons cited by respondents were the lack of such help, stigmatisation, distrust of specialists, and the negative experiences of their acquaintances who had sought such help. News feeds often feature reports of human rights violations in mental health institutions in Ukraine, including in foreign media [223, 224, 225, 226, 227, 228]:

*«When I started talking about patients’ right to legal protection, I heard: «Don’t you understand that they’re all schizophrenics?» — Larysa Savitska, legal aid office volunteer [224]*

Specialists from different countries propose strategies for managing pregabalin withdrawal syndrome and treating patients with mental and behavioural disorders resulting from pregabalin use, and report on successful cases [8, 181, 229]. With this in mind, it is recommended to improve the mental health care system and develop and implement protocols for the treatment of disorders associated with pregabalin use, drawing on the existing experience of countries around the world.

Of particular note is the report on the effectiveness of harm reduction programmes as a tool for preventing psychoactive substance use and the negative consequences of such use. Researchers from France report on the effectiveness of standards for the evaluation and selection of prevention programmes in the field of psychoactive substance use using ASPIRE standards, and the standards themselves operate with concepts that are universal for most Euro-cultural contexts. In addition, standards for ensuring access to them are provided to interested parties free of charge, including translation [230].

A striking example of the effectiveness of harm reduction programmes is Portugal, where in 2001 (at the height of the opioid epidemic) the possession of psychoactive substances was decriminalised and a harm reduction policy was introduced. This led to a sharp decline in substance use and addiction, HIV infection, overdose deaths, crime, and stigma associated with substance use [231].

## CONCLUSIONS

Pregabalin is popular among psychoactive substance users in the information space, but most of them are unaware of the risks of such use and combination with other psychoactive substances. Respondents who participated in the study formed the impression that pregabalin was safe after trying it, which subsequently led to systematic use, negative consequences, and an inability to stop taking it.

Pregabalin is available for recreational use in Ukraine, including to minors, despite prescription restrictions on the sale of medicines containing it. Respondents noted the availability of medicines containing pregabalin in the pharmacy network, the possibility of purchasing them without a prescription in regular pharmacies, and, if necessary, obtaining a prescription without medical indications for its use in healthcare facilities.

The patterns of recreational use of pregabalin by respondents, taking into account the dosages and duration of use, are dangerous in terms of the development of dependence, withdrawal syndrome, mental and physical health disorders, negative social consequences, accidents, and material losses. Respondents indicated that maximum daily doses ranged from 1,500 to 9,000 mg, which significantly exceeded the dose taken when they first used the drug (150 to 750 mg).

Pregabalin was used as a tool for chemsex by both women and men. This is due to its psychoactive effects and meets the requirements that a substance must have for use for this purpose. As a result, respondents reported an increase in the number of sexual partners and casual sexual encounters, as well as the emergence of new sexual desires and experiences. An alarming “signal” according to the study is the respondents’ disregard, under the influence of pregabalin, for measures to reduce the harm from risky sexual behaviour, which exacerbates the burden of infectious and non-infectious diseases.

The study found that respondents had difficulty giving up recreational use of pregabalin and showed low levels of commitment to seeking help from the existing mental health care system for substance users. Among the reasons cited by respondents were: lack of necessary medical and psychological care, stigmatisation of mental disorders and substance use, negative experiences of seeking such care, distrust of medical staff (especially psychiatrists) and lack of need.

## RECOMMENDATIONS

Educating young people about the risks and negative consequences of recreational use of pregabalin is relevant and appropriate, given the popularity of pregabalin among users of psychoactive substances and young people, social networks and art, low public awareness of the negative consequences of recreational use of pregabalin and its combination with other psychoactive substances, the existence of dangerous patterns of recreational use of pregabalin in terms of the development of mental and physical health disorders, and negative socio-economic consequences. This approach is recommended by a number of foreign researchers [100, 192].

It is advisable to conduct educational activities among medical students and practising doctors regarding the risks of prescribing pregabalin and the adequate assessment of the benefits/harms of such a prescription, given the tendency of doctors to prescribe pregabalin, including off-label, and the absence of warnings in the Ukrainian medical information space about the addictive potential of pregabalin and the dangers of combining it with other drugs. To strengthen the promotion of countermeasures against the recreational use of pregabalin, it is proposed to review/update/develop training programmes for medical professionals at the undergraduate and postgraduate levels, as well as to offer training courses/seminars for doctors, pharmacists and psychologists in continuing professional development on the risks of pregabalin misuse, signs of dependence and first aid for intoxication. The need to inform, first and foremost, doctors who prescribe pregabalin about the management (including risks) of this drug is confirmed by data from foreign researchers [96, 158, 232], and measures to raise awareness among doctors are recommended by the National Health Service of Scotland [233] and the Canadian Medical Association [234]. Data from Alshahrani et al. also emphasise the advisability of raising pharmacists’ awareness of the abuse of psychotropic drugs and the consequences of dispensing them without a prescription [99].

The implementation/adaptation of international protocols and strategies for the prevention of psychoactive substance use in the Ukrainian context of recreational pregabalin use, using ASPIRE standards that have proven effective in the prevention of psychoactive substance use, make sense in most Euro-cultural contexts and are distributed free of charge [230].

Consider the possibility of conducting and financing scientific research using quantitative methods and including data collection on recreational use of pregabalin in the national monitoring of the drug and alcohol situation in Ukraine. Based on the results of scientific research, amend the Procedure for Monitoring the Drug and Alcohol Situation in Ukraine, approved by Resolution of the Cabinet of Ministers of Ukraine No. 689 of 10 July 2019 “Issues of monitoring the drug and alcohol situation in Ukraine”, taking into account the lack of quantitative data on the recreational use of pregabalin. According to Duarte, N et al., state monitoring of trends in the use of psychotropic drugs is key to preventing uncontrolled dispensing and related deaths from overdose [81].

Review and amend the Risk Management Plans for medicines containing pregabalin, indicating the danger of abuse and dependence on pregabalin, the development of withdrawal syndrome and the combination of pregabalin with other medicines.

Develop standards for the provision of medical care for mental and behavioural disorders resulting from the use of pregabalin, taking into account the existence in Ukraine of dangerous patterns of recreational use of pregabalin in terms of the development of dependence, withdrawal syndrome, mental and physical health disorders, the occurrence of emergency conditions and requests for emergency and planned medical care in the context of pregabalin use, and the lack of standards for the provision of medical care for such conditions and disorders.

Expand the capacity of public health and healthcare service providers to promote and conduct educational work on risky sexual behaviour and the distribution of harm reduction measures among pregabalin users, taking into account the use of pregabalin as a tool for chemsex, an increase in the number of sexual partners, the emergence of new sexual desires and experiences after starting recreational use of pregabalin, and neglect of harm reduction measures for risky sexual behaviour while using pregabalin.

Consider the possibility of introducing specialised harm reduction programmes for users of psychoactive substances, including pregabalin. The effectiveness of harm reduction programmes is demonstrated by the example of Portugal, where in 2001, at the height of the opioid epidemic, the possession of psychoactive substances was decriminalised and a harm reduction policy was introduced, after which the country saw a sharp decline in the use of psychoactive substances and addiction, the spread of HIV infection, overdose mortality, crime, and stigma associated with psychoactive substance use [231].

Create conditions for free and anonymous treatment by including mental health care in the outpatient and inpatient medical care package and providing such care for mental disorders, including mental and behavioural disorders due to psychoactive substance use, taking into account the barriers to seeking medical care due to the stigma associated with mental disorders and psychoactive substance use, as well as mistrust of professionals in terms of maintaining the confidentiality of treatment.

Improve the mental health system by preventing human rights violations in mental health institutions in order to provide the population with access to quality psychological, social and medical care aimed at preventing self-medication with pregabalin.

## AUTHOR CONTRIBUTIONS

Mykyta Lapin: research concept and design, data collection and analysis, interpretation of results.

Maryna Shevchenko: critical review, final approval of the article.

## USE OF ARTIFICIAL INTELLIGENCE

The authors confirm that no artificial intelligence-based technologies were used in the writing and editing of this article.

## Data Availability

All data produced in the present study are available upon reasonable request to the authors

## APPENDIX A: PROFILE OF RECREATIONAL USE OF PSYCHOACTIVE SUBSTANCES BY RESPONDENTS

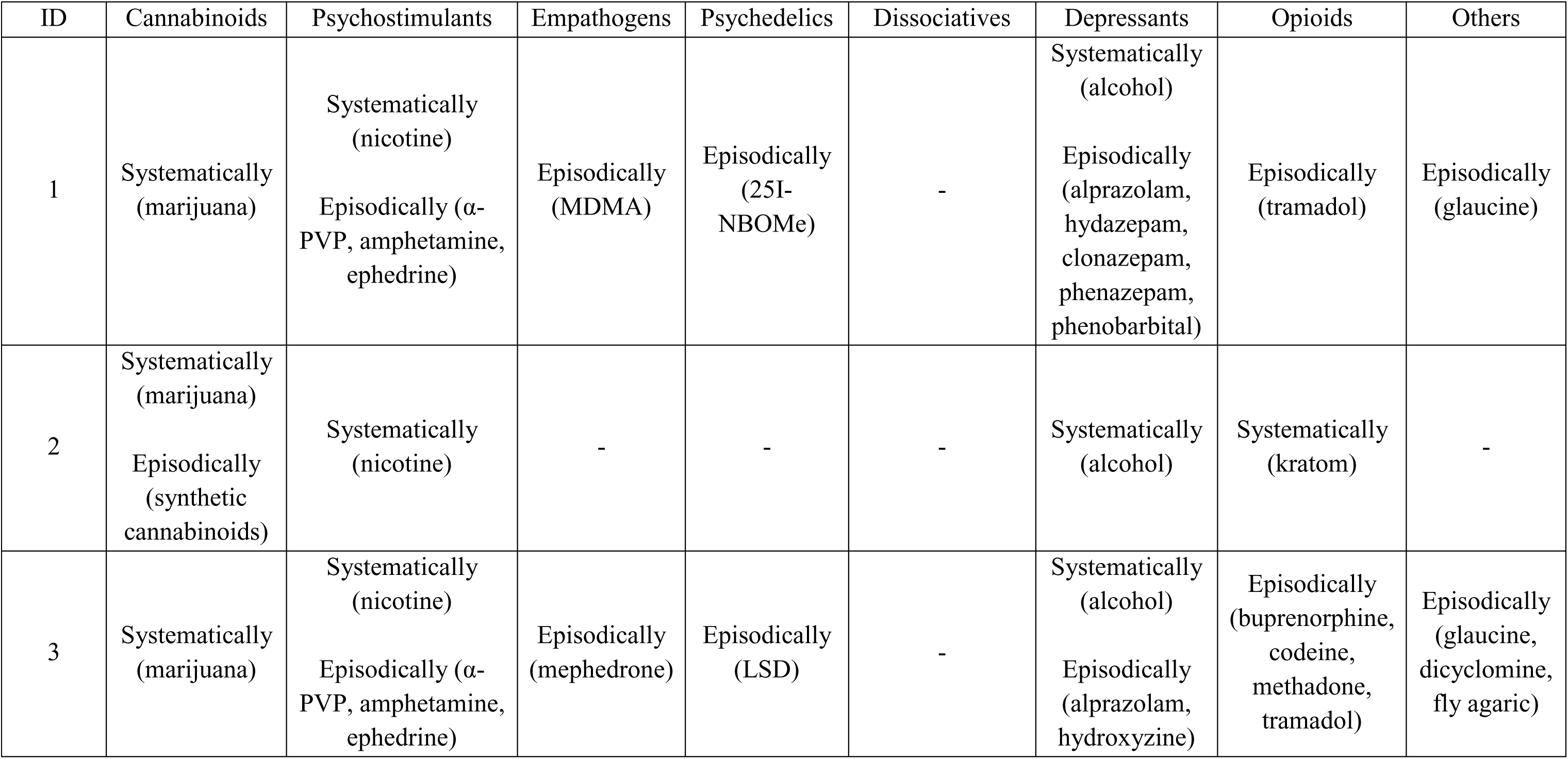
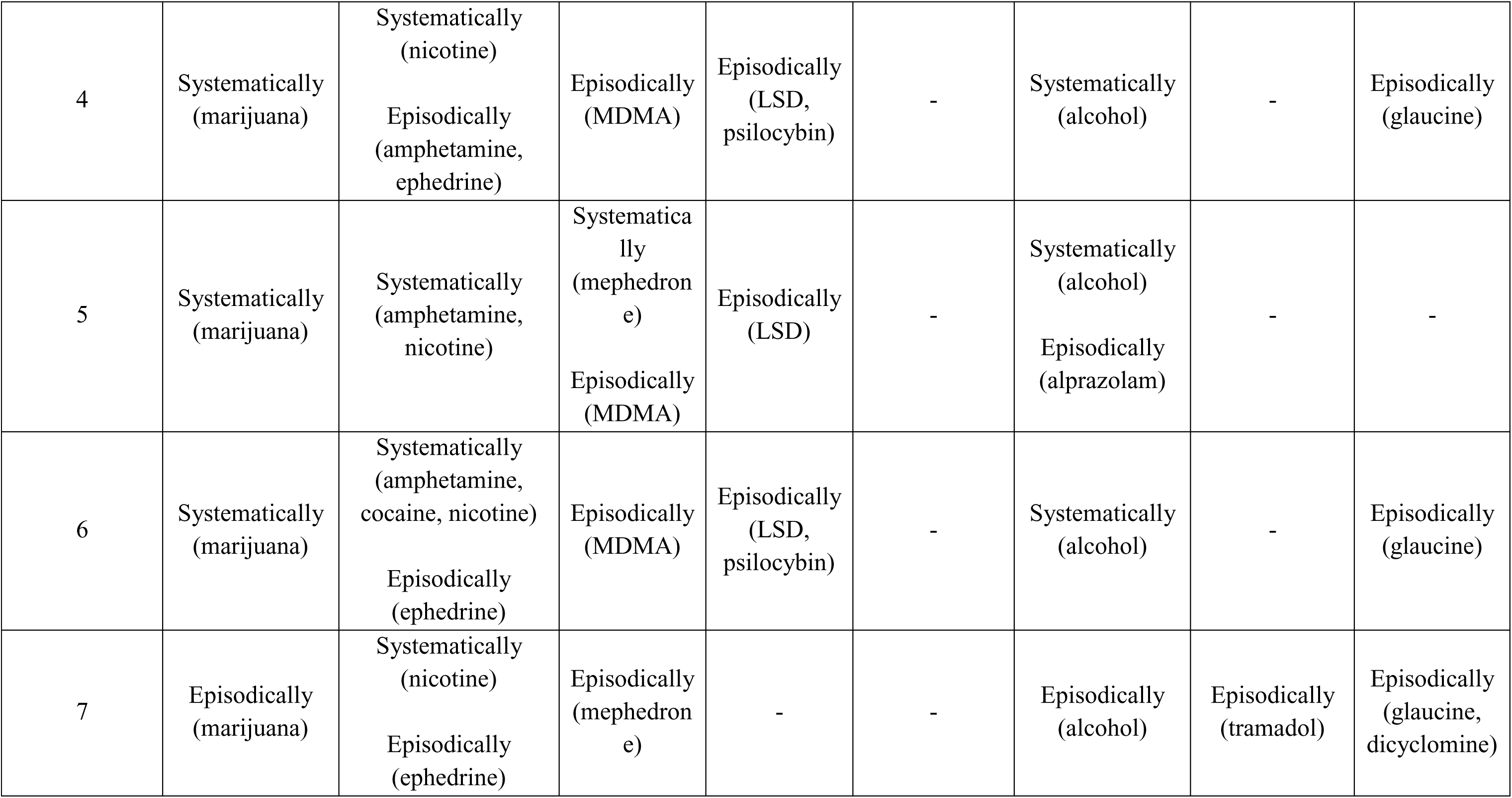
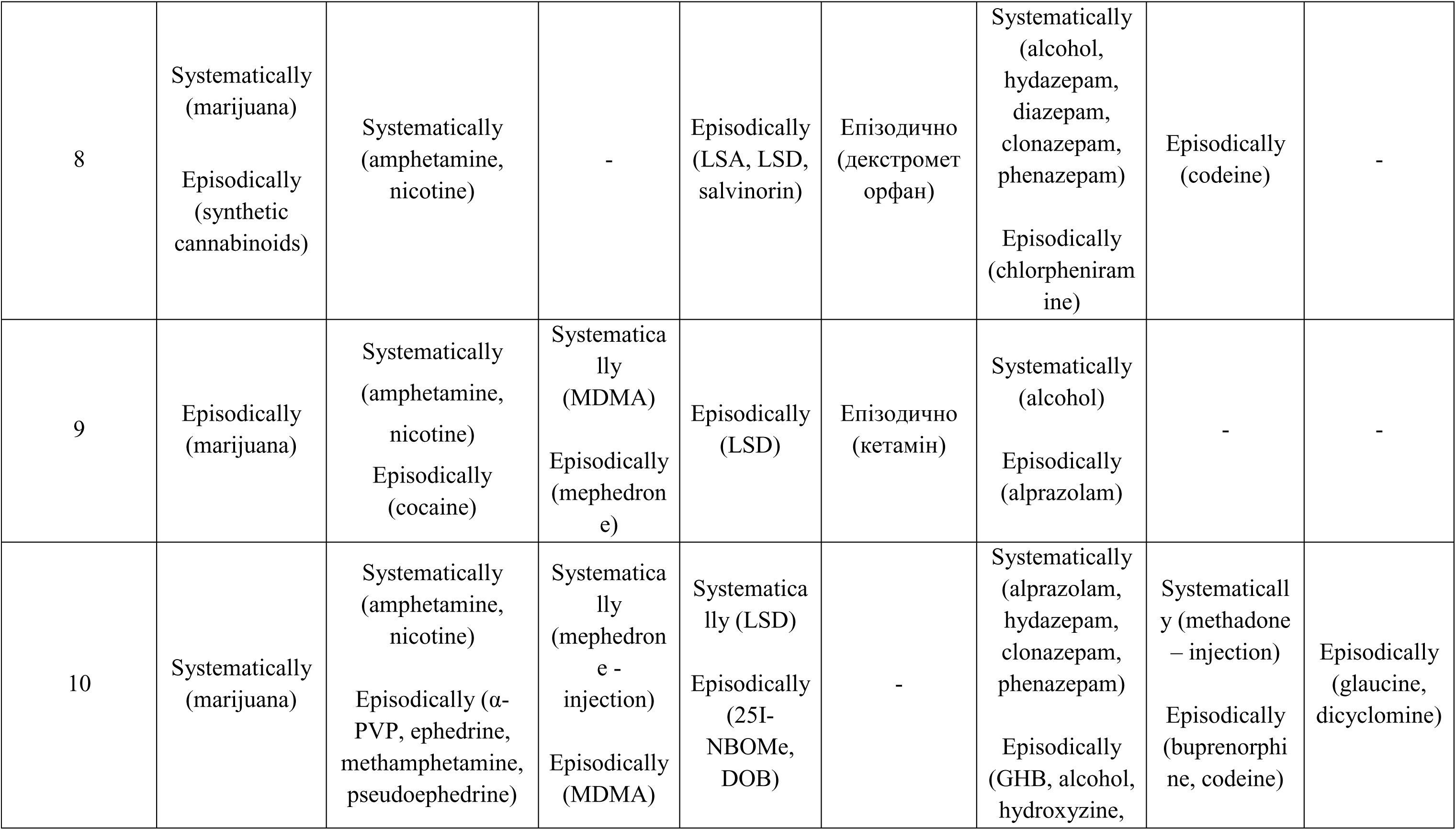
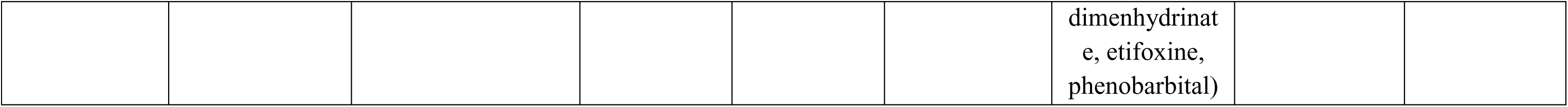

## APPENDIX B: COMBINATION OF PREGABALIN WITH OTHER PSYCHOACTIVE SUBSTANCES

APPENDIX B COMBINATION OF PREGABALIN WITH OTHER PSYCHOACTIVE SUBSTANCES
| ID | Cannabinoids | Psychostimulants | Empathogens | Psychedelics | Dissociatives | Depressants | Opioids | Others |
| --- | --- | --- | --- | --- | --- | --- | --- | --- |
| 1 | Marijuana | Nicotine | - | - | - | Alcohol | - | - |
| 2 | Marijuana | Nicotine | - | - | - | Alcohol | Kratom | - |
| 3 | Marijuana | Amphetamine,<br>ephedrine, nicotine | - | - | - | Alcohol,<br>alprazolam,<br>hydroxyzine | Codeine,<br>tramadol | Glaucine,<br>dicyclomine |
| 4 | Marijuana | Amphetamine,<br>ephedrine, caffeine,<br>nicotine | - | - | - | Alcohol | - | Glaucine |
| 5 | Marijuana | Nicotine | - | - | - | Alcohol | - | - |
| 6 | - | Ephedrine, nicotine | - | - | - | Alcohol | - | Glaucine |
| 7 | - | Ephedrine, caffeine,<br>nicotine | - | - | - | Alcohol | Tramadol | Glaucine,<br>dicyclomine |
| 8 | - | Caffeine, nicotine | - | - | - | Alcohol,<br>hydazepam | - | - |
| 9 | - | Amphetamine,<br>nicotine | - | - | - | Alcohol | - | - |
| 10 | Marijuana | Nicotine | MDMA | - | - | - | Codeine,<br>methadone | Glaucine |

## ACRONYMS

α-PVP: alpha-pyrrolidinopentiophenone
DALY: disability-adjusted life year
FDA: Food and Drug Administration
GABA: gamma-aminobutyric acid
GHB: gamma-hydroxybutyric acid
HIV: human immunodeficiency viruses
LSA: lysergic acid amide
LSD: lysergic acid diethylamide
MDMA: 3,4-methylenedioxymethamphetamine

